# Gene-environment interplay in internalising and externalising psychopathology in adolescence

**DOI:** 10.1101/2025.04.02.25325078

**Authors:** Agnieszka Musial, Andrea G. Allegrini, Rosa Cheesman, Angelica Ronald, Essi Viding, Thalia C. Eley, Kaili Rimfeld, Robert Plomin, Margherita Malanchini

## Abstract

A combination of genetic and environmental factors working in interplay is thought to underlie differences in symptoms of psychopathology between adolescents. Yet, studies that have investigated gene-environment interaction in isolated aspects of developmental psychopathology lack robust effects, highlighting the need for a more comprehensive approach. We adopted a multivariable framework to investigate gene-environment interaction in internalising and externalising symptoms of psychopathology in a sample of 3,337 16-year-olds from the Twins Early Development Study. We used penalised regression models to examine the main effects of genetic factors (G), indexed combining 13 polygenic scores for psychopathology, and environmental factors (E), measured by combining multiple environmental exposures during childhood and adolescence, on symptoms of psychopathology. We also examined their additive effects (G+E) and their interaction (G×E). Polygenic scores accounted for, on average, 2.7% of the variance in symptoms of psychopathology, with stronger predictions for externalising symptoms, while environmental measures alone accounted for an average of 7.1% of the variance. G+E accounted for an average of 9.1% of differences between adolescents in symptoms of psychopathology. We observed small G×E effects for internalising symptoms, accounting for an average of 1.1% of the variance. Children with a higher genetic risk showed higher levels of internalising symptoms, especially when exposed to more chaos at home and harsher parenting. Our findings indicate that genetic and environmental influences contribute additively, underscoring the importance of jointly considering both factors to enhance our understanding of youth psychopathology. At the same time, our results highlight the persistent challenges involved in identifying robust G×E effects.

## Introduction

Symptoms of psychopathology include abnormalities in behavioural, cognitive, and adaptive functioning. These symptoms often emerge in childhood, leading to an increased risk of poor educational, socio-economic, and health outcomes ^1–3^. In the current work, we focus on two broad domains of psychopathology: internalising and externalising problems ^4^. Internalising problems include symptoms of anxiety and mood disorders (e.g., depression), they typically onset between ages 6 and 21 and affect an estimated 6.5% of children and adolescents worldwide ^2,5^. Externalising problems can manifest as behavioural problems or impulse-control deficits (e.g., symptoms of attention-deficit hyperactivity disorder (ADHD) and conduct disorder), often begin between ages 7 and 15 and have a global prevalence of 5.7% ^2,5^. These disorders and their symptoms can persist throughout the lifespan and have been found to predict adult mental illness up to two decades later ^6^.

Studies investigating the origins of internalising and externalising symptoms of psychopathology in adolescence have found evidence for the contribution of genetic factors. Twin studies estimated that genetic differences accounted for between 37 and 67% of differences between adolescents in anxiety and depression, depending on whether symptoms were reported by adolescents or their parents ^7^, and between 36 and 78% of differences between adolescents in ADHD symptoms and conduct problems ^8–11^. DNA-based approaches to estimating heritability– i.e., the proportion of variation in a trait accounted for by variation in single nucleotide polymorphisms (SNPs), provide lower heritability estimates than those reported by twin studies. SNP heritability in adolescent samples has been estimated at 5% for anxiety, less than 0.001% for depression ^7^ and 0-22% for ADHD ^7,12^ and 5-13% for conduct problems in adolescence and early adulthood ^7,13^. Genetic effects on adolescent psychopathology have also been estimated using polygenic scores, which aggregate hundreds of SNP associations emerging from gene-discovery studies into a single composite index ^14,15^. Polygenic scores for psychopathology and neurodevelopmental disorders have been found to explain up to 5% of the variance in childhood and adolescent behavioural and emotional problems ^16–18^.

In addition to genetic effects, environmental factors have been linked to symptoms of internalising and externalising psychopathology ^19–21^. Shared environmental factors– those environmental factors that contribute to similarities between family members ^22^–have been found to play a moderate role in symptoms of depression and anxiety, with estimates ranging between 10-30% ^23^, but only a limited role in the development of externalising symptoms. On the contrary, nonshared environmental factors– those environmental experiences that are specific to each individual in a family– have been found to play a substantial role across internalising and externalising symptoms, accounting for 40- 59% of the variance in anxiety ^24^, 36-55% in depression ^25^, 30% in ADHD ^9,26^ and 37% in conduct problems ^26^.

Despite the substantial contribution of nonshared environmental effects to variation in internalising and externalising symptoms, studies have struggled to identify the specific environments that constitute nonshared environmental influences. In the hope of bridging this *missing nonshared environment gap* ^27^, studies have combined multiple environmental measures ^28^. However, these poly-environmental scores constructed using environmental measures collected across development, such as differential parenting and classroom environment, were found to account for only up to 4.7% of the nonshared environmental variance symptoms of behavioural problems in adulthood ^24^. Interestingly, poly-environmental composites explained 19% of the total variance in symptoms of behaviour problems in adolescence ^24^, suggesting that these scores do not mainly operate via the nonshared environment and do not solely work as environmental exposures, suggesting gene-environment interplay.

An interplay between genetic and environmental factors has been proposed as a key mechanism for internalising and externalising symptoms of psychopathology ^29^. Two main forms of gene-environment interplay have been described: gene-environment correlation and gene-environment interaction ^30–33^. Gene-environment correlation refers to how individuals experience environments that are in line with their genetic dispositions, which statistically is reflected in the covariance between an individual’s genotype and environmental factors ^31^. For example, children with a higher genetic risk for antisocial behaviour were found to elicit harsher punishment from their parents, if compared to children with a lower genetic risk ^34^. Gene-environment interaction (G×E) describes the moderating effect of genetic propensities on environmental exposures. Gene-environment interaction occurs when individuals experience and respond to the environment differently depending on their genetic dispositions ^35^.

The diathesis-stress framework describes how genetic dispositions interact with environmental factors to influence behavioural outcomes. According to this model, individuals with a genetic vulnerability (the diathesis) are particularly susceptible to the effects of environmental adversity, which may increase their risk for developing psychopathology ^36^. Therefore, genetic influences can either amplify or mitigate the impact of environmental factors, leading to different psychological and behavioural outcomes. Conversely, high-risk environments overshadow genetic dispositions, while low-risk environments enable genetic differences to manifest ^37^. Alternative models, for example the differential susceptibility theory ^38,39^ propose that, due to higher genetic sensitivity, certain individuals might be more responsive to their environmental conditions, whether positive or negative. Unlike the diathesis-stress model, which primarily emphasizes vulnerability to adversity, the differential susceptibility framework highlights the potential for enhanced development in supportive environments as well. However, this model is yet to be supported by robust empirical evidence ^37^. In line with the diathesis stress model, early twin research found support for the role of G**×**E in internalising symptoms, where adolescents genetically predisposed to depression may exhibit increased sensitivity to developing symptoms when exposed to social adversity ^40^, as well as in externalising symptoms, for example, a study found that children responded differently to childhood maltreatment and differences in parenting styles, based in part on their genetic disposition towards conduct disorder ^41,42^.

The advent of polygenic scores has led to an increase in research investigating G×E in symptoms of psychopathology beyond twin samples ^17,43^. A recent investigation found that genetic susceptibility to major depressive disorder increased the risk of depressive symptoms in adolescents who were exposed to higher levels of criticism from their parents ^44^. Another study found that a composite measure of early environmental risk moderated the association between the ADHD polygenic score and externalising symptoms in adolescence, nonetheless, these interaction effects were weak, accounting for up to 0.4% of the variance ^17^. Several other studies investigating G×E in symptoms of psychopathology did not find evidence for gene-environment interaction. For example, a higher genetic risk for ADHD was not found to interact with childhood maltreatment in predicting symptoms of ADHD in adolescence ^45^. Similarly, childhood maltreatment was not found to moderate the association between a higher genetic disposition for risky behaviour and externalising problems in adolescence ^46^.

A shift in methodology, moving beyond piecemeal approaches that consider specific genetic and environmental variables in isolation is likely to lead to breakthroughs in the discovery of the G×E effects underlying internalising and externalising symptoms of psychopathology. Particularly, machine learning approaches offer the opportunity of integrating a wide range of predictors and therefore comprehensively examine the role of genetic and environmental factors and their interaction. Potential gains in prediction might be achieved by combining polygenic scores for several psychiatric disorders. This approach leverages the observed genetic correlation between different traits, which implies that to the extent that the genetic signal between two traits overlaps, a polygenic score derived from one trait can predict variance in another trait ^47^. The accuracy of this prediction depends on the extent of the shared genetic signal between the traits. Combining multiple polygenic scores was found to strengthen the polygenic prediction of cognitive, anthropometric and health-related traits ^47^ and of symptoms of behaviour problems in youth ^48^. Further gains might be achieved by combining multiple environmental measures and estimating their additive effects in predicting symptoms of psychopathology and their interaction (G×E) ^49^. This multivariable approach resulted in greater prediction accuracy of academic achievement in adolescence ^50^.

The current study builds on this multivariable framework and applies machine learning methods to comprehensively investigate G×E in symptoms of internalising and externalising psychopathology in adolescence, leveraging the wealth of environmental, behavioural and psychological data available in the Twins Early Developmental Study (TEDS). We capture symptoms of internalising by considering measures of anxiety and depression and externalising across behaviour problems, ADHD, and conduct disorder, rated by 16-year-olds and their parents. We measure genetic disposition towards psychopathology by combining thirteen polygenic scores derived from well-powered genome-wide association studies (GWAS) of psychopathology and neurodevelopmental disorders; and combine multiple environmental measures, including measures of the family, home and school environment collected at ages 9, 12 and 16.

First, we investigate the main effects of genetic (G) and environmental (E) variables on externalising and internalising symptoms in adolescence. Second, we examine their additive effects (G+E); and third, their interaction (G×E). Lastly, we consider gene-environment correlation alongside G×E. Through gene-environment correlation, individuals may seek or create environments that align with their genetic propensities, which introduces a layer of complexity in interpreting G×E effects ^43,51,52^. As such, considering gene-environment correlation is crucial to the accurate interpretation and identification of G×E effects ^53,54^.

## Results

### Genetic prediction of internalising and externalising symptoms of psychopathology (G models)

Figure 1 presents the variance in symptoms of psychopathology predicted additively by the 13 selected polygenic scores as well as the regression coefficients for the effect of each polygenic score on parent-reported and self-reported symptoms of psychopathology at age 16. Full results and fit parameters are presented in Supplementary Table 1. On average, polygenic scores additively predicted 2.4% of the variance in parent-rated and 3.0% of the variance in self-rated symptoms of psychopathology. For parent-rated data, the prediction ranged from 3.1% for symptoms of ADHD to 1.9% for anxiety and callous-unemotional traits. For self-reported symptoms, polygenic scores explained between 4.3% of the variance in ADHD symptoms and 1.6% of the variance in symptoms of depression. Models were more predictive of externalising than internalising symptoms (3.2% vs 1.9% of the variance). Polygenic scores constructed from the GWAS of externalising and ADHD were the strongest genetic predictors of behaviour problems while the neuroticism polygenic score evinced the strongest prediction when considering internalising symptoms of anxiety and depression.

**Figure 1.**
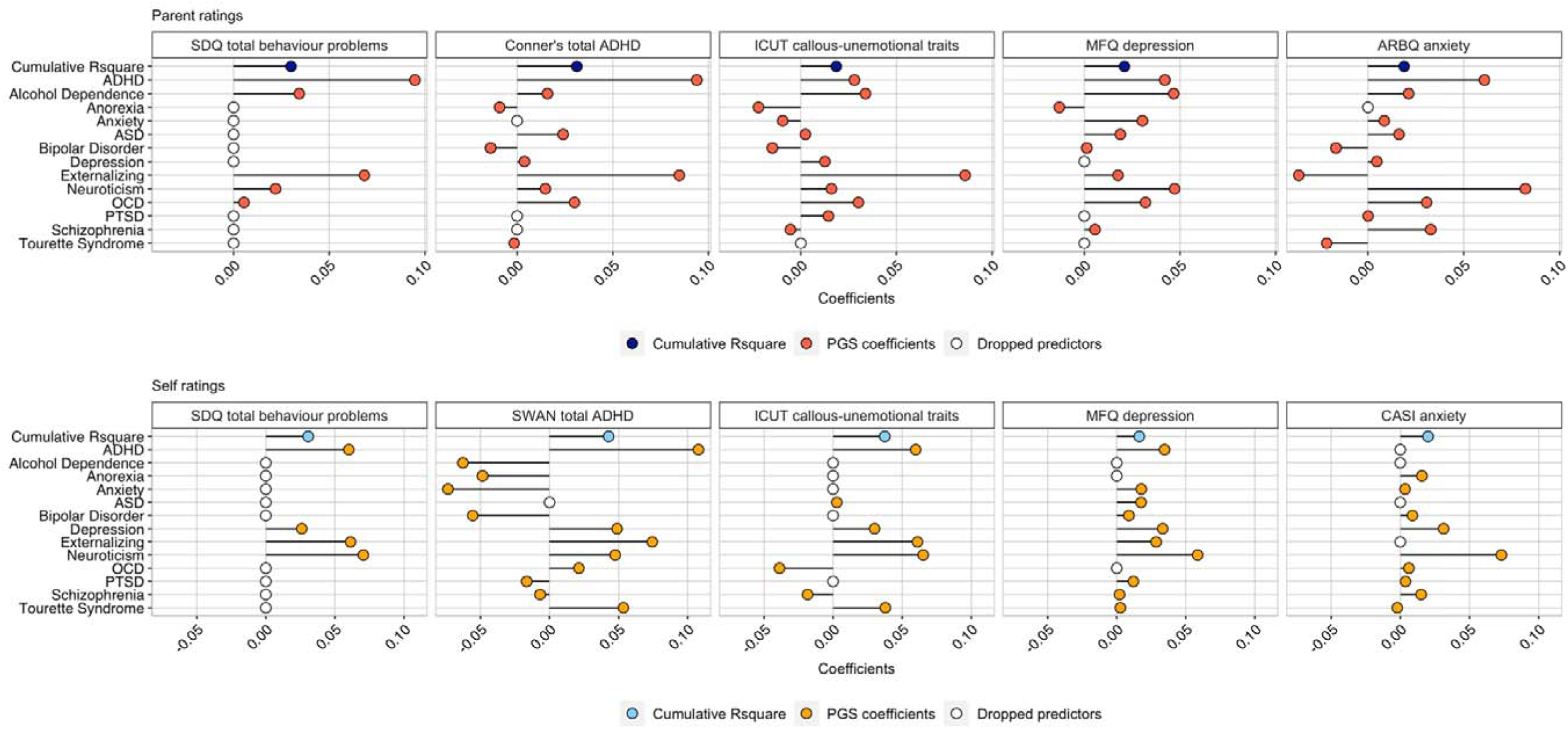
Results of the G models predicting internalising and externalising symptoms of psychopathology at age 16 combining 13 polygenic scores. The first row of each panel represents the cumulative proportion of variance explained in symptoms of psychopathology (Rsquare). The remaining rows represent elastic net coefficients of the association between each of the polygenic scores and measures of psychopathology. Dropped predictors indicate variables with 0 coefficients that were dropped from the elastic net model.

### Environmental prediction of internalising and externalising symptoms of psychopathology (E models)

Figure 2 depicts the extent of the prediction of symptoms of internalising and externalising psychopathology from environmental exposures measured in childhood and adolescence (ages 9, 12 and 16). Full results and fit parameters are presented in Supplementary Table 2. On average, environmental measures accounted for a similar proportion of variance in internalising and externalising symptoms of psychopathology rated by parents and self-reported by the adolescents (7.7% and 6.5%, respectively).

**Figure 2.**
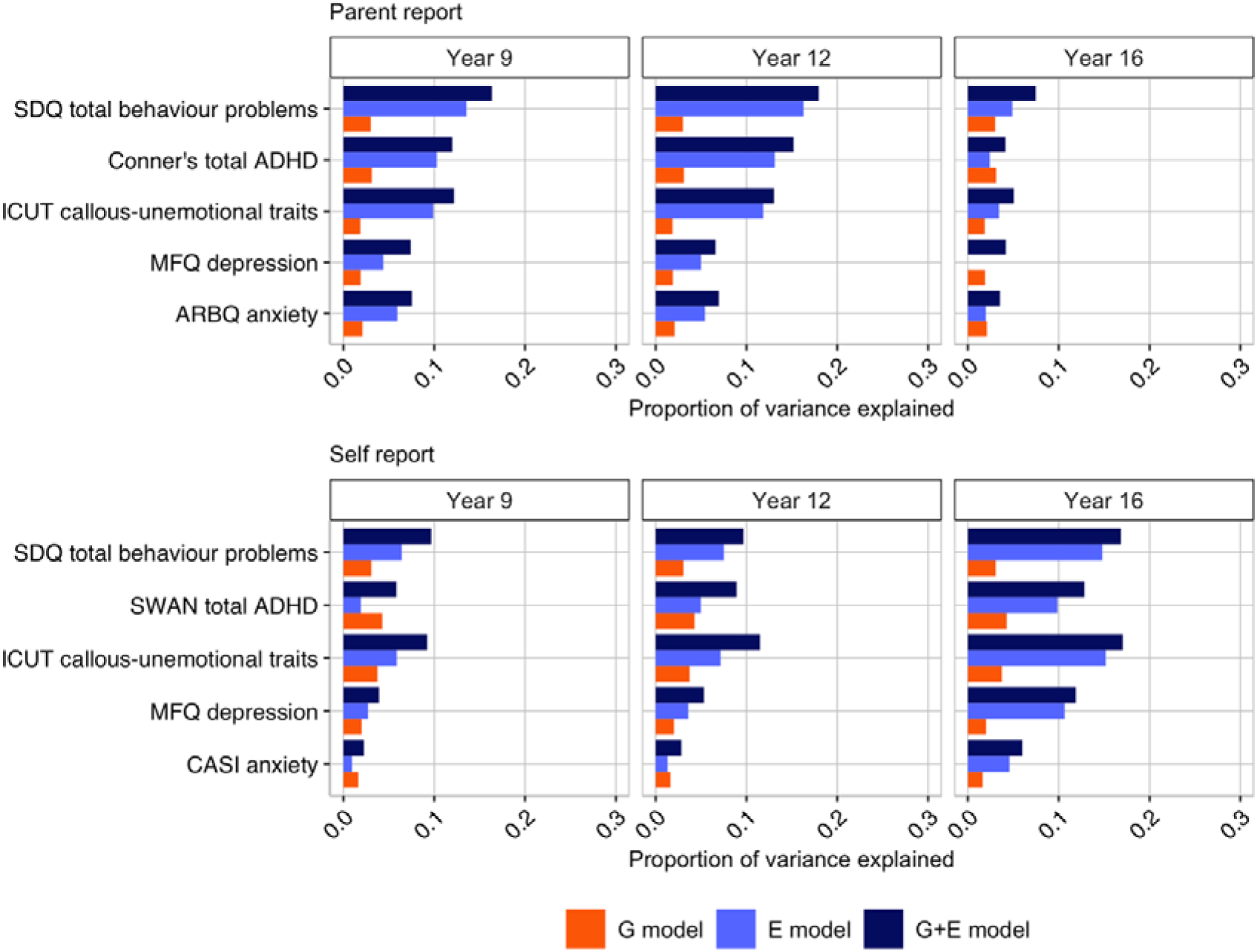
Variance in symptoms of internalising and externalising psychopathology additively accounted for by the polygenic scores and environments measured at ages 9, 12 and 16, compared to the variance explained by G and E models. The top bars indicate the additive prediction of additively combined G and E effects, the middle bars indicate the proportion of variance explained by environmental measures and the bottom bars indicate the proportion of variance explained by the polygenic scores.

Although the overall prediction was comparable, different environmental measures contributed to the prediction of symptoms across raters. When considering parent-rated measures of psychopathology, more variance was accounted for by the parent-rated environments measured in childhood and early adolescence (i.e., 8.8% at age 9 and 10.3% at age 12), as compared to parent-rated environments measured at age 16 (3.2%). The opposite was observed when we considered self-rated symptoms of psychopathology, as the variance accounted for by self-rated environmental measures collected at age 16 was twice that of ages 9 and 12 (11.0% vs 3.6% and 4.9%, respectively). Parent-rated environmental factors were more predictive of externalising symptoms (9.5% on average) than they were of internalising symptoms (4.5% on average). Similar pattern emerged for self-rated environments, which were more predictive of externalising (8.0%), rather than internalising symptoms (4.0%). The most predictive environmental factors of parent-rated symptoms of psychopathology were aspects of the parental feelings (e.g., feelings of anger or frustration) and chaos scales (e.g., noise or disorganisation) and life events measured when the children were 9 years old (e.g., financial difficulties or the birth of a new sibling) (Supplementary Figure 1). Child-reported parental feelings and home chaos were the most predictive measures of self-reported symptoms of psychopathology throughout adolescence (Supplementary Figure 2).

### Bringing together genetic and environmental predictors (G+E)

Figure 2 shows the proportion of variance in parent and self-rated symptoms of psychopathology additively accounted for by genetic and environmental measures at ages 9, 12 and 16. G and E effects largely combined additively, as including both G and E in the model improved the prediction of psychopathology symptoms. Polygenic scores and parent-rated environmental factors age 9 additively accounted for 11.1% of the variance in parent-rated symptoms. Combining parent-rated environments measured at age 12 and polygenic scores accounted for 11.9% of the variance in symptoms of psychopathology at age 16, while polygenic scores + parent-rated environments at age 16 accounted for 4.9 % of the variance in symptoms of psychopathology at the same age. For self-rated environmental measures, the variance explained additively was 6.4% for age 9 environments, 7.6% for age 12 environments and 12.9% for age 16 environments. The largest differences in the proportion of variance explained between E and G+E models (quantified as a delta Rsquare) were observed for depression at age 9 (delta Rsquare= 3.0%), ADHD traits at age 12 (delta Rsquare= 2.1%) and behaviour problems at age 16 (delta Rsquare= 2.6%) for parent-rated data and for ADHD traits at ages 9, 12 and 16 (delta Rsquare= 3.9%, 3.9% and 3.0%, respectively) for self-rated data. The differences in prediction between E and G+E models were similar when looking across raters, with a mean delta Rsquare of 2.6% at age 9, 2.2% at age 12 and 1.9% at age 16. Across parent and self-rated measures, the mean delta Rsquare was greater for externalising, 2.3% at age 9, 1.7% at age 12 and 2.0% at age 16, than for internalising symptoms of psychopathology, 2.1% at age 9, 1.6% at age 12 and 1.4%% at age 16. Full results and fit parameters of the E models are presented in Supplementary Table 3. Results of the sensitivity G+E models that were conducted using the total genotyped sample, including dizygotic co-twins are presented in Supplementary Table 4. Additionally, Supplementary Tables 5, 6 and 7 show results of G+E models for cross-rater and sex-specific prediction, respectively. Comparison of the proportion of variance explained by sensitivity models is presented in Supplementary Figure 3.

### Integrating gene-environment interaction effects (G×E)

We detected a total of 27 significant interactions between the polygenic scores and environments measured at ages 9, 12 and 16 in predicting symptoms of internalising psychopathology such as depression and anxiety (Figure 3). Supplementary Figures 4-6 plots the G×E that emerged as significant in the continuous analysis for twins selected in the +/- 1 standard deviation quadrants of G and E. Individuals scoring high on G consistently tend to exhibit more behaviour problems than the low-G group. Likewise, the high-E group showed more behaviour problems compared to the low-E group. The majority of the interactions are ordinal, where genetic effects remain in the same direction but vary in strength across environments; however, several showed a disordinal pattern, in which the environmental measures of home environment and parenting influenced the development of internalising psychopathology in opposite ways depending on one’s genetic risk for schizophrenia or bipolar disorder.

**Figure 3.**
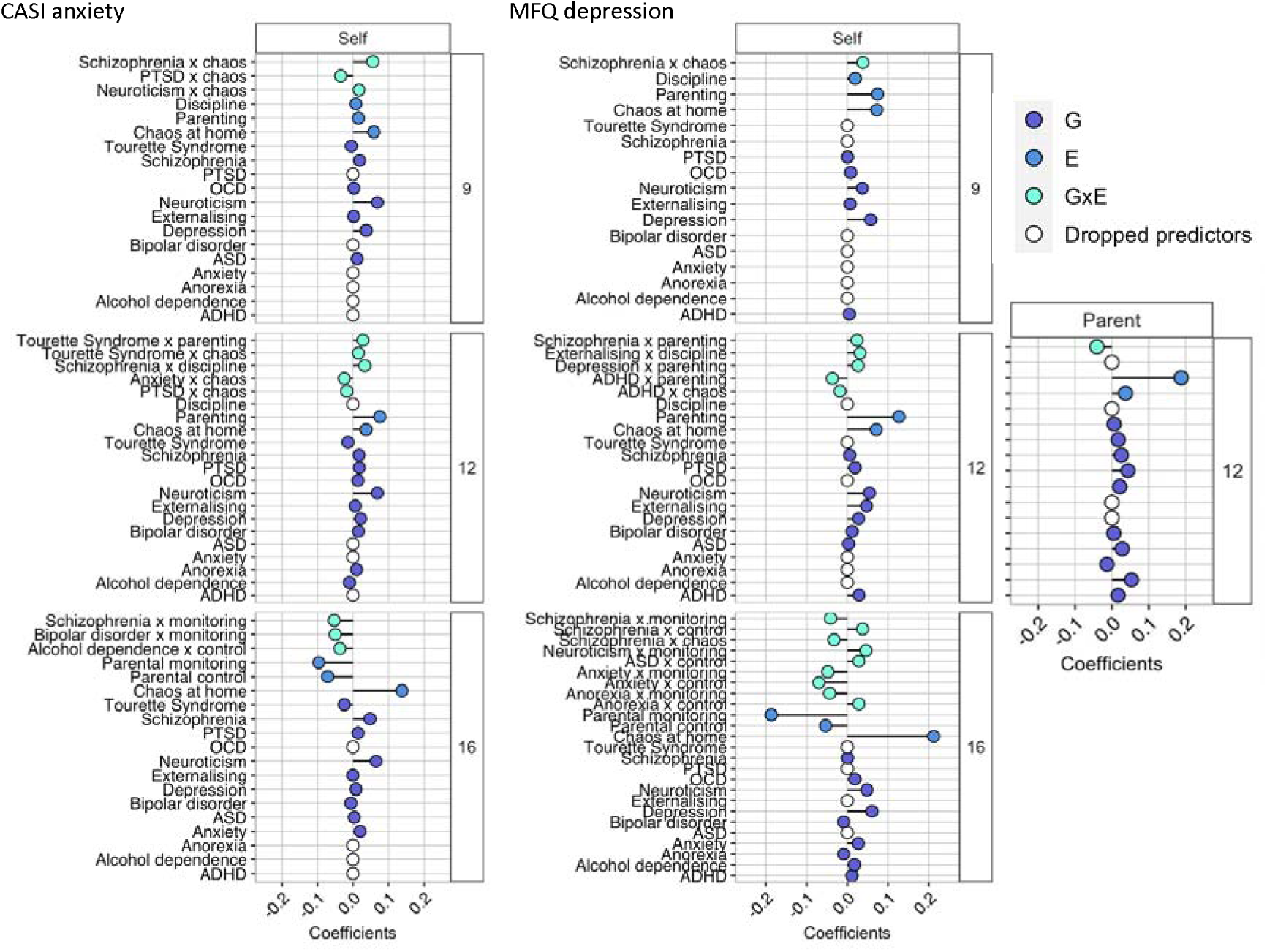
Coefficients of the additive G+E+G×E models predicting a measures of depression and anxiety at age 16 using genome-wide polygenic scores (G), environmental measures (E) and interactions between the polygenic scores and environments (G×E). Rows represent elastic net coefficients of the association between G, E, G×E and self-reported CASI anxiety (left panel) and self- and parent-reported MFQ depression (right panel). Dropped predictors indicate variables with 0 coefficients that were dropped from the elastic net model.

When considering environmental measures at age 9, the interactions that remained significant involved: (1) genetic disposition to neuroticism and schizophrenia interacting with self-reported chaos at home in predicting symptoms of anxiety, and (2) between the schizophrenia polygenic score and self-reported chaos at home in prediction of depression symptoms. These finding suggests the occurrence of ordinal G×E, where genetic risk for neuroticism and schizophrenia exacerbates vulnerability to anxiety and depression in the presence of a chaotic home environment. When considering environmental measures at age 12, ordinal interactions were detected between polygenic scores for anxiety, schizophrenia, depression and externalising and environmental measures of self-reported home chaos, discipline and parenting in prediction of internalising psychopathology. A crossover interaction was observed between genetic risk for schizophrenia and parent-reported depressive symptoms, where at low levels of parental discipline, higher genetic risk was positively associated with depressive symptoms, but at high levels of discipline, the relationship reversed, with higher genetic risk being negatively associated with depressive symptoms.

When considering environments measured contemporaneously at age 16,we observed a mixture of ordinal and crossover interactions. The association between genetic vulnerability to depression, bipolar disorder and schizophrenia and anxiety symptoms were moderated by higher levels of parental control and monitoring. Similarly, parental control and monitoring moderated the association between polygenic scores for anxiety and schizophrenia and symptoms of depression. The crossover interactions involved genetic risk for bipolar disorder and parental monitoring in predicting anxiety symptoms as well as the schizophrenia polygenic score and measures of chaos and parental control predicting depressive symptoms. Those with a higher genetic risk for bipolar disorder exhibited more anxiety symptoms under high parental monitoring but fewer under low monitoring, while those with higher schizophrenia polygenic score showed more depressive symptoms in structured, controlled environments but fewer in chaotic settings. Adding the identified interaction terms to the previously estimated G+E models resulted in a mean delta Rsquare of 0.7% for age 9 environments, 0.8% for age 12 environments and 1.9% for age 16 environments (Supplementary Table 8). Regression weights for all predictors in the G+E+G×E models for CASI anxiety and MFQ depression are presented in Figure 3.

### Tests of gene-environment correlation

Because G and E indices are unlikely to be purely genetic and environmental and correlations between them can create spurious G×E, we additionally performed tests of rGE. Correlations between predicted values from G and E elastic models are presented in Supplementary Figure 7 for parent- and self-reported data. Based on these correlations being generally less than 0.20 for parent-reported data and polygenic scores and less than 0.15 for self-reported environmental data and polygenic scores, we did not observe substantial rGE effects in our sample. We have subsequently run mediation models of rGE for those phenotypes for which significant G×E was detected. We tested for the mutual mediation of the values predicted from the G and E models of age 9, 12 and 16 environments on symptoms of internalising and externalising psychopathology, namely MFQ depression and CASI anxiety. Based on the Bonferroni corrected p-value of p<0.004, significant environmentally-mediated effects were observed for MFQ depression using environment measured at ages 9, 12 and 16 and for CASI anxiety using environments measured at age 16. Significant genetically-confounded effects were observed for MFQ depression using age 12 and 16 environments and for CASI anxiety using age 12 environments (Supplementary Table 9). Environmental mediation effects increased with age, ranging from 5.3– 14.9% at age 12 to 14.5–27.5% by age 16, with the highest mediation observed for self-reported MFQ depression in late adolescence. In contrast, genetically confounded effects were smaller (4.1–9.4%) and remained relatively stable across ages, peaking at 6.6% for self-reported MFQ depression at age 12. These findings suggest a growing influence of environmental factors on adolescent mental health over time. For these G and E indices we also observed the significant interaction effects, making it difficult to disentangle the extent to which rGE between these variables affected the result of the G×E analyses, even though the elastic net model used to estimate the variance explained by G, E and their interaction controlled for multicollinearity between predictors.

## Discussion

The present study applied a comprehensive framework to examine the extent to which genetic and environmental factors predict internalising and externalising symptoms of psychopathology in adolescence. Our main goal was to investigate gene-environment interaction in symptoms of psychopathology moving beyond piecemeal approaches that had considered only a few variables in isolation to integrate a wide range of predictors by leveraging machine learning approaches. The wealth of developmental data collected as part of TEDS provided us with the ideal setting to test the effectiveness of these comprehensive models. The results point to the importance of considering both genetic and environmental measures. Yet, we found that genetic and environmental predictors mostly combined additively, and that evidence for their interplay, particularly in terms of interaction effects, was modest.

We first explored the extent to which our multivariate genetic instrument considering genetic vulnerability across several neurodevelopmental disorders and psychopathologies could predict differences between adolescents in internalising and externalising symptoms. We found that, collectively, polygenic scores accounted for a small proportion of the variance (2.7% on average). The strongest predictors of most psychopathology outcomes were polygenic scores that captured genetic dispositions towards externalising behaviour and ADHD. Our results are in line with previous findings that the polygenic scores for psychiatric disorders that are currently available can more reliably capture individual differences in externalising symptoms than internalising symptoms, particularly in developmental samples ^16,17,81,82^. One potential explanation is differences in statistical power. Gene-discovery studies that have identified genetic variants related to externalising symptoms have been generally more powerful, with sample sizes reaching 1.5 million individuals ^83^, as compared to less than 400 thousand individuals included in the most powerful gene-discovery study of internalising symptoms, that of neuroticism ^84^. Additionally, reporting challenges related to internalising symptoms in children—such as difficulties with self-awareness or the ability to articulate emotions—might contribute to the weaker associations observed between polygenic scores and internalising symptoms during adolescence ^85^. However, whilst the polygenic score prediction was overall weaker, we found that symptoms of anxiety and depression were better captured by genetic disposition towards neuroticism than externalising.

Measured environmental factors, which included multiple aspects of the home environment, parenting and life events, accounted for a modest proportion of variance in symptoms of psychopathology in adolescence (on average 7.7% in parent-reported and 6.5% in self-reported symptoms, respectively). Differences in prediction accuracy could be observed depending on who reported on environmental experiences. Parent-rated environmental exposures measured earlier in development when the children were 9 and 12 years old, were better predictors of adolescent psychopathology than environmental exposures rated by parents at age 16. On the other hand, environmental exposures self-rated by the children at age 16 were more accurate predictors of self-rated symptoms of psychopathology at 16 years old, if compared to earlier self-reported environmental exposures. This pattern is consistent with developmental expectations. In early childhood, children are highly dependent on their caregivers, and peer relationships have not yet become central. As a result, parenting and the home environment are likely to exert a stronger influence during this period. By age 16, however, adolescents typically experience greater independence, and their own perceptions of their environment may become more relevant to their emotional and behavioural functioning. This shift reflects the growing autonomy and identity formation characteristic of adolescence ^86–88^. These findings highlight the complexity of measuring environmental influences across development and the potential influence of reporter bias, reliability, and sensitivity at different time points.

Among the measured environmental factors, positive and negative parental feelings and the perception of a chaotic home environment were identified as the most important predictors of psychopathology symptoms, both when rated by parents and youths. The fact that parental feelings, particularly feelings of frustration and impatience, emerged as important factors associated with the development of behavioural and emotional problems in adolescence highlights the significance of parental emotional regulation and communication in contributing to adolescents’ emotional well-being. A chaotic home environment as measured by the chaos scale, which encompasses disorganization, lack of routine, and unpredictability in the household environment, might provoke stress and feelings of instability in children and adolescents, potentially contributing to the development of behavioural problems and emotional distress. However, these associations may be confounded by the effects of genes, as environmental measures often show substantial genetic influence ^89^, and that the use of polygenic scores as G indices in the current study is an imperfect method of controlling for G. These results are nevertheless in line with the findings of longitudinal studies, which demonstrated that negative aspects of parenting are associated with the development of antisocial behaviour in childhood and adolescence and highlighted the role of both genetic and environmental effects ^90–92^.

Our results also highlight how considering genetic and environmental factors in combination led to more accurate predictions of symptoms of psychopathology in adolescence. Integrating genetic information to solely environmental predictions contributed an additional 2% of the variance on average. Despite the small effect sizes associated with our individual genetic instruments, we found significant G×E effects in the prediction of internalising psychopathology, specifically symptoms of depression and anxiety. At age 9, genetic risk for schizophrenia and neuroticism interacted ordinally with home chaos, predicting greater symptoms of self-reported anxiety and depression. This pattern of interaction supports a diathesis-stress model, indicating that individuals with higher genetic vulnerability experience increasingly worse mental health outcomes under adverse conditions, while those with a lower genetic vulnerability are less affected by chaos at home. Similar ordinal interactions emerged from our analyses of environmental exposures at age 12: children with a higher genetic vulnerability to anxiety, schizophrenia, and depression were at an increased risk of developing internalising symptoms at age 16 when exposed to unpredictable or harsh environments at age 12.

However, a distinct interaction pattern was observed for genetic risk for ADHD, where higher genetic risk was associated with fewer depressive symptoms in chaotic or unstructured environments. This suggests that a disposition towards ADHD may confer adaptive advantages in less structured settings, potentially due to increased stimulation or reduced pressure to conform to rigid structures, therefore in line with what proposed by the differential susceptibility hypothesis ^38,39^.

At age 16, genetic vulnerability to depression, anxiety, schizophrenia, and bipolar disorder interacted with higher levels of parental control and monitoring, in predicting higher levels of internalising symptoms. Rather than serving as a protective factor, strict parental control appeared to exacerbate the effect of genetic risk on internalising symptoms. Exceedingly high levels of parental control might increase stress, restrict actual or perceived autonomy, or create an environment of heightened scrutiny that might intensify symptoms of anxiety and depression, particularly during adolescence, a time of discovery and exploration ^93^. Crossover interactions indicate that genetic risk does not simply predict worse outcomes but rather heightens sensitivity to environmental conditions in both positive and negative ways. Specifically, individuals with a higher schizophrenia polygenic score **e**xhibited fewer depressive symptoms in chaotic environments but more symptoms in structured, controlled settings, whereas those with a higher bipolar disorder polygenic score showed greater anxiety symptoms under high parental monitoring but reduced symptoms when monitoring was low.

It is important to acknowledge that the detection of several G×E effects in internalising symptoms does not necessarily indicate the prevalent presence of G×E in adolescent psychopathology. Further research and replication studies are needed to establish the robustness and generalizability of these observed interaction effects. Our findings of a lack of robust and widespread G×E effects on individual differences in symptoms of psychopathology are in line with what has been previously reported for academic achievement using this same multivariable approach ^49,94^. Notably, several of the detected interactions between the polygenic scores and environmental measures also exhibited significant indirect effects in genetic correlation analyses, highlighting the need to interpret G×E findings in light of gene-environment correlation effects ^53,54^. Significant environmentally-mediated effects were found for depression and anxiety . The identified rGE effects make it challenging to interpret the G×E effects because it is difficult to separate how much of the observed G×E interaction is due to G×E itself versus the rGE, even though the elastic net model was used to control for multicollinearity between genetic, environmental, and interaction predictors.

Historically, it has been difficult to pinpoint the specific genetic and environmental factors that work in interaction to influence symptoms of psychopathology, particularly when considering robust replicable effects ^37,95^. While it could be that these interactive effects are genuinely small, hence with the current sample size we are unable to differentiate true signal from noise ^49^, studies that have been successful in detecting more robust and widespread G×E effects have mostly used broader, all-encompassing measures of environmental exposures, such as differences between schools or boarder political and social contexts ^96,97^. For instance, a multilevel modelling study found that latent differences between schools interact with children’s genetic predisposition to ADHD. Children with higher risk for ADHD showed greater differences in achievement depending on what school they went to, with higher-performing schools compensating for genetic risk ^97^. On the contrary, studies that attempted to examine more defined, individual and subjective environmental effects under closer scrutiny, including the present investigation, have found little evidence for the influence of G×E on a wide array of behavioural outcomes ^37,49,98,99^.

Several factors could also contribute to a lack of widespread, substantial interaction effects. First, the limited predictive power of psychiatric polygenic scores that only partly summarise genetic vulnerability to symptoms of psychopathology. Second, our study’s statistical power, although adequate for detecting substantial G×E effects exceeding 1% of the variance, was insufficient to pinpoint more subtle effects ^17,100^. While smaller G×E effects might not be easily detectable with our sample size, they are valuable for understanding the pathways between genes and environments ^101^. Although larger G×E effects might be of more immediate practical utility ^17,102^.

Third, incorporating variables that are more closely linked to biological processes, for example sleep patterns and cortisol levels, could provide a deeper understanding of the interplay between psychological and physiological factors ^105–107^. Another potential avenue to be explored in relation to identifying G×E effects could incorporate investigation of epigenetic signatures, including DNA methylation algorithms capturing accelerated pace of biological aging as unbiased measures of environmental exposure to stress ^28,108–110^. Fourth, these very small G×E effects are likely to be even smaller and underpowered when studied across non-European samples due to the lower prediction of genetic instruments across different ancestries ^114^. This leads to difficulties in generalising findings across populations and ancestries ^115,116^, further contributing to inequalities in the discovery of evidence-based targets for intervention and prevention strategies ^117^. Fifth, a further potential limitation of this study is the lack of inclusion of age and sex in the interaction models, as these factors may moderate the developmental trajectories of psychopathology.

In conclusion, our findings highlight the importance of considering both genetic and environmental factors when studying internalising and externalising symptoms of psychopathology in adolescence. While these factors mainly combined additively, we observed widespread yet small G×E associated with differences between adolescents in internalising symptoms, in line with extant literature.

Although small, these interaction effects highlight how a uniform approach to prevention and interventions might not suit all adolescents, as their reactions to environmental exposures vary depending in part on their genetic propensities ^37,118,119^.

## Method

Analyses for this project were preregistered with the Open Science Framework (OSF) (https://osf.io/dzqnu/). The hypotheses and deviations from the pre-registration are listed in Supplementary Note 1. Analytic scripts are available on the OSF page and https://github.com/CoDEresearchlab/GE-interplay-developmental-psychopathology.

### Sample

Our sample included twins born in England and Wales between 1994 and 1996 enrolled in the Twins Early Development Study (TEDS). For a detailed description of the sample and its representativeness, see Supplementary Note 2, Supplementary Table 10 and ^55^. In the present study we investigated the prediction of symptoms of internalising and externalising psychopathology at age 16, using the polygenic scores and measures of the environment collected when the twins were approximately 9, 12 and 16 years old in order to explore the effects of previous and contemporaneous environments. DNA was collected from a subsample of 7,026 unrelated twins and 3,320 dizygotic twin pairs. The genotyping procedure involved two different platforms (AffymetrixGeneChip 6.0 and Illumina HumanOmniExpressExome-8v1.2) and was conducted in two separate waves. For details on genotyping, imputation and quality control, see ^56^.

Our models were tested using data from unrelated twins, which was achieved by randomly selecting one dizygotic twin per each genotyped pair. The resulting sample size ranged from 3,337 (56% females) to 640 (63% females) individuals with developmentally complete phenotype data for G and E models. Details on sample size per model are provided in Supplementary Tables 1-7. While the statistical power of the present study to detect interaction effects below a 1% threshold may be limited, the primary objective is to examine these interactions collectively within a multivariable framework, akin to a variance components approach. This method enables a comprehensive assessment of interaction effects by considering them within a broader, integrated model, potentially enhancing our understanding of their collective influence on developmental outcomes ^57^. We also conducted sensitivity analyses using the total genotyped sample, i.e., including the dizygotic co-twins.

### Measures

Variables in the current study included polygenic scores (G), parent and self-rated measures of the environment at age 9, 12 and 16 (E), as well as parent and self-rated symptoms of internalising and externalising psychopathology measured at age 16. Figure 4 provides a visual illustration of G, E and psychopathology variables used in these analyses.

**Figure 4.**
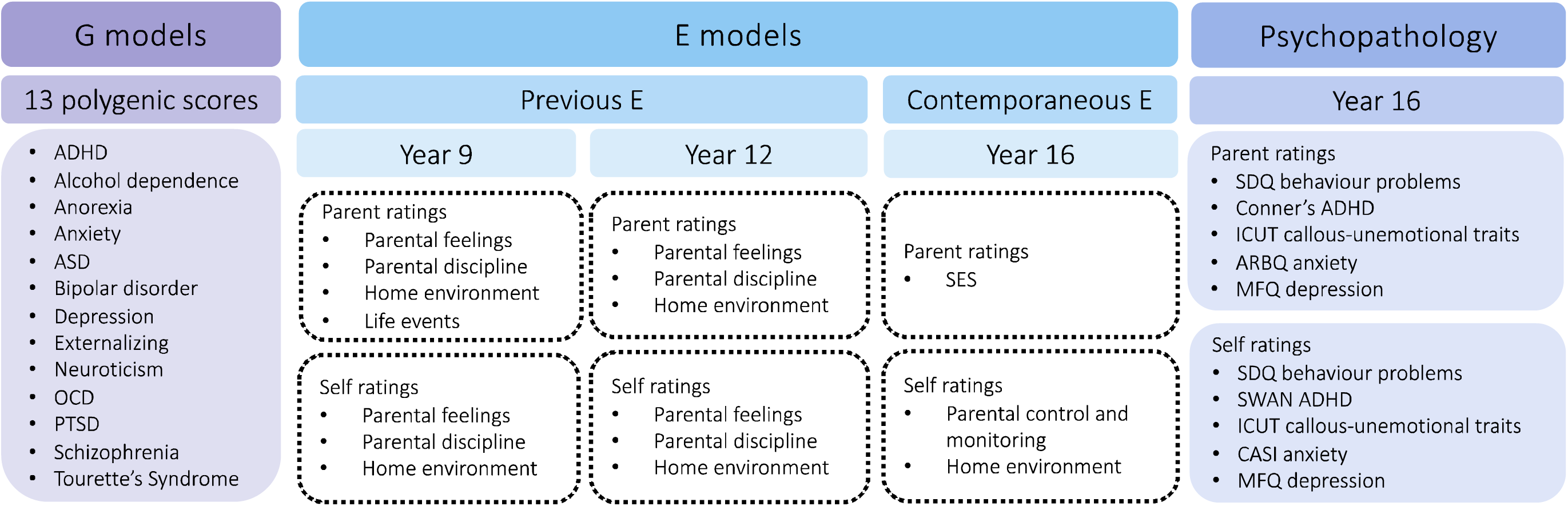
A visual representation of the genetic, environmental and psychopathology variables used in the current study. G models refer to predicting symptoms of psychopathology using polygenic scores. E models refer to predicting symptoms of psychopathology using environment data collected at ages 9, 12 and 16.

### Polygenic scores

We used polygenic scores derived from the 11 most powerful GWAS of neurodevelopmental disorders and broadly defined psychopathology from ^58^. In addition, we used polygenic scores derived from the GWAS of neuroticism ^59^ and externalising behaviour ^60^. The GWAS from which these 13 polygenic scores have been derived are listed in Supplementary Table 11. Polygenic scores were constructed in LDpred2-auto, using the UK Biobank reference panel and SNPs restricted to HapMap3+ variants (for details, refer to Supplementary Note 3) ^61^.

### Environmental measures

Environmental predictors included parent and self-rated measures of the environment assessed at ages 9, 12 and 16, that had been previously found to predictive behavioural and emotional problems in adolescence ^24^. At age 9, we included measures of parental feelings ^62^, discipline ^63^, home environment ^64^ and life events. At age 12, the measures included all those used at age 9, except for life events. At age 16, the measures included a composite measure of the family socioeconomic status (SES), control and monitoring ^65^ and home environment ^64^. These environmental variables are presented in Figure 4 and described in greater detail in Supplementary Table 12. More details on each measure can be found in the TEDS data dictionary, available at the following link: https://datadictionary.teds.ac.uk/home.htm

### Symptoms of internalising and externalising psychopathology

We investigated the prediction of internalising and externalising aspects of psychopathology, such as behaviour problems at age 16, measured using the Strengths and Difficulties Questionnaire (SDQ) ^66^. Additional measures of externalising psychopathology included assessment tools of ADHD, such as the Conners Rating Scale ^67^ and the Strengths and Weaknesses of Attention-Deficit/Hyperactivity Symptoms and Normal Behaviors (SWAN) ^68^ and conduct problems, such as the Inventory of Callous-Unemotional Traits (ICUT) ^69^. Measures of internalising psychopathology included the Anxiety-Related Behaviours Questionnaire (ARBQ) ^70^, Childhood Anxiety Sensitivity Index (CASI) ^71^ and measures of mood disorders, such as the Mood and Feelings Questionnaire to assess depressive symptomatology (MFQ) ^72^. These measures are described in greater detail in Supplementary Table 13 and the TEDS data dictionary (https://datadictionary.teds.ac.uk/home.htm).

### Analyses

#### Data pre-processing

All environmental variables were residualised for age and sex to mitigate the results being in part driven by age- or sex-related variations. Polygenic scores were additionally regressed on the first 10 genetic principal components and genotyping chip. The obtained standardised residuals were used in all analyses. Environmental variables and measures of psychopathology were assessed for normality and square root or inverse transformations were applied for variables with skewedness larger than 1 or lower than -1. For details on transformation methods used, distributions and correlations between transformed and raw variables, see Supplementary Note 6, Supplementary Table 14 and Supplementary Figures 8-10.

#### Gene and environment (GE) prediction

We estimated the independent (G and E) and additive (G+E) effects of the 13 polygenic scores and environmental measures using a shrinkage model referred to as elastic net regularization to overcome problems of multicollinearity and overfitting ^50,73,74^. We performed the elastic net analyses using the R package glmnet ^74^ implemented in caret ^75,76^. For every model tested, we performed the 10-fold nested repeated cross-validation repeated 100 times, using nestedcv for R ^77,78^.The additive effect of the polygenic scores and environmental variables was estimated by fitting all G and E predictors together in elastic net models for each psychopathology phenotype and observing the additional variance explained. For a detailed description of the method, see Supplementary Note 5 and ^49^.

#### Gene-environment interaction (G×E)

To identify interactions between the polygenic scores and environmental measures (G×E), we employed a hierarchical lasso procedure implemented in the R package *glinternet* (group-lasso interaction network) ^76,79^. This procedure helps minimise the impact of multiple testing and maximise power to detect small G×E effects ^79^. To select interactions, *glinternet* uses a group lasso and performs variable selection on groups of variables, simultaneously eliminating or maintaining them in the model. In addition, *glinternet* was used in this instance rather than elastic net approach because for an interaction between two variables to be selected, the main effects of both these variables need to be detected based on non-zero model coefficients ^79^. The selected 2x2 interactions between the polygenic scores and environments were reintroduced to the G+E models and changes in prediction accuracy (i.e., variance explained) were examined.

#### Sensitivity analyses

In addition to the models involving the unrelated sample, we fitted the G+E models for the total sample, that is including dizygotic co-twins, as well as samples of males and females. We also tested cross-rater prediction, estimating the variance explained in parent-reported psychopathology using self-reported environmental data and vice versa.

#### Gene-environment correlation (rGE)

In addition to testing for G×E, we also performed a set of exploratory analyses to investigate the role of rGE in internalising and externalising psychopathology and account for the correlation between genetic and environmental factors on G×E effects ^43,51^. We modelled rGE effects in two ways. First, we correlated the predicted values from G and E elastic net models and estimated the overlap between G and E effects on symptoms of psychopathology ^50^. Correlating the predicted G and E values would reflect the degree of overlapping information between genetic and environmental factors influencing symptoms of psychopathology that can conceptually be interpreted as indication of rGE ^50^.

Second, we used mediation models (Supplementary Note 6 and Supplementary Figure 11) to test whether the indirect effects of G on symptoms of psychopathology were mediated by E and, vice versa, whether the effects of E were mediated by G ^50^. In the mediation models, we used the predicted values from G and E elastic net models to assess the degree to which the effects of polygenic scores and environmental influences on variation in symptoms of psychopathology are mutually mediated. The schematic representation of rGE mediation models are presented in Supplementary Figure 11.

The mediation analyses were performed using *lavaan* for R ^76,80^.

## Code availability

Analytic scripts are available on the OSF page and https://github.com/CoDEresearchlab/GE-interplay-developmental-psychopathology.

## Acknowledgements

We gratefully acknowledge the ongoing contribution of the participants in the Twins Early Development Study (TEDS) and their families. TEDS is supported by the UK Medical Research Council (MR/V012878/1 and previously MR/M021475/1. This study was funded by a starting grant awarded to MM by the School of Biological and Behavioural Sciences at Queen Mary, University of London. MM is supported by a Jacobs Foundation Research Fellowship that funds AM postdoctoral fellowship. RC is supported by the Jacobs Foundation (#2023-1510-00), the European Research Council (#101115949, (#101045526), and the Research Council of Norway.

## Author information

AM and MM conceived and designed the study. AM analysed the data. MM supervised the work. AM and MM wrote the paper with helpful contributions from AGA, RC, AR, EV, TCE, KR and RP. All authors contributed to the interpretation of the data, provided critical feedback on paper drafts and approved the final draft.

