## Supplementary Material for "Gene-environment interplay in internalising and externalising psychopathology in adolescence"

### Supplementary Notes

#### Supplementary Note 1: Hypotheses pre-registered with the Open Science Framework (OSF) and deviations from the pre-registration.

The following hypotheses were preregistered with the Open Science Framework (OSF) (<https://osf.io/dzqnu/>):

Hypothesis 1: Overall, environmental measures will predict more variance in developmental psychopathology symptoms than polygenic scores.

Hypothesis 2: The proportion of variance in symptoms of developmental psychopathology explained by G×E will be modest, generally less than 1%.

Hypothesis 3: Effects are likely to be stronger when the same person (parent or child) rates the environment and symptoms of developmental psychopathology as compared to cross-rater analyses.

Hypothesis 4: Stronger prediction will be achieved for externalising (ADHD, conduct problems), rather than internalising (anxiety, mood disorders) measures of developmental psychopathology.

Hypothesis 5: Effects will not differ substantially between males and females.

Although we expected to see differences in patterns of results across measures and raters, we focused on comparing the magnitude effect sizes (Rsquare), rather than statistical significance due to power limitations of the current sample. While large enough to detect effects of polygenic scores and environmental measures accounting for 1% of the variance with 80% power, according to power calculations performed by ^1^, G×E effects explaining 0.1% of the variance require tens of thousands of individuals to be detected ^2^.

In the pre-registration we specified 14 polygenic scores but in the paper we have decided to drop the hypomania polygenic score due to poor predictive power. We also decided to focus on total symptom scores for both psychopathology and environmental data, that meant dropping the symptom-specific sub-scales and individual items (such as peer relationship problems) and only using total scales in our models (such as total behaviour problems).

#### Supplementary Note 2: Description of the TEDS sample.

Our sampling frame consisted of up to 3,337 unrelated twins born in England and Wales between 1994 and 1996 who have been enrolled in the Twins Early Development Study (TEDS) ^3^. The TEDS twins have been assessed a dozen times from infancy through early adulthood on a wide range of behavioural, psychological, cognitive, physical and environmental measures ^3^. Data collection procedures included questionnaires administered by post, by telephone and online, as described in an overview of TEDS ^3^. Details can be found in the TEDS data dictionary: <https://www.teds.ac.uk/datadictionary/home.htm>). The sample of TEDS twins is representative of the UK population in terms of ethnicity and socioeconomic status (SES) (Supplementary Table 1); for details of representativeness and attrition, see ^4^. Individuals with severe medical conditions were excluded from analyses. These conditions include detrimental prenatal and postnatal conditions, as well as other conditions that could seriously impact later development. In addition, twins with uncertain and unknown zygosity were excluded from the analyses. Zygosity was recorded using a parent questionnaire of physical similarity between twins, with 95% accuracy when ascertained by DNA tests ^5^.

#### Supplementary Note 3: Construction of the polygenic scores.

Polygenic scores were calculated as the weighted sums of each individual’s genotype across all single nucleotide polymorphisms (SNPs):

$$\mathrm{PGS}_{\mathrm{ki}}=\sum_{j=i}^{m} \hat{\beta}_{\mathrm{kj}} g_{\mathrm{kji}}$$

Where PGS_ki_ represents the individual i’s polygenic score based on summary statistics from GWAS_k_. $\hat{\beta}_{\mathrm{kj}}$is an estimate of marker j’s effect size for discovery trait k, that is, the effect of having one copy of the reference allele at SNP_kj_ . g_kji_ is individual i’s genotype at marker j for discovery trait k, coded as having either 0, 1 or 2 copies of the reference allele at marker kj.

We used polygenic scores constructed using LD-pred2-auto ^6^ with infinitesimal prior, which corrects for local linkage disequilibrium (LD; i.e. correlations between SNPs). We used the UK Biobank samples as a reference panels for the LD structure and restricted to HapMap3+ variants.

#### Supplementary Note 4: Details of elastic net regularization.

We estimated the independent (G and E) and joint (G+E) prediction of the 13 polygenic scores and environmental measures using a shrinkage model referred to as elastic net regularization to overcome problems of multicollinearity and overfitting ^7^. Elastic net regularization tries to minimise the following loss function ^8^:

||y − X𝛽||2 + λ(α*|β|1 + (1−α)*|β|2)

where ||y–X’β||2 is the residual sum of squares, |β|2 is the sum of the squared betas (the L2 penalty), |β|1 is the sum of the absolute betas (the L1 penalty) and X is an N*P (‘N’ observations and ‘P’ predictors) matrix of polygenic scores and environmental measures (for details, see ^9^).

For every model tested, we performed the nested repeated cross-validation, using nestedcv for R ^10,11^. The nested repeated cross-validation splits the data into inner and outer folds. In the inner fold, performed the 10-fold cross-validation repeated 100 times to select the model that minimises the Root Mean Square Error (RMSE), which indicates the smallest cross-validation error ^12^. We then fitted the model on this inner fold and tested the model on the hold-out outer fold, followed by a final cross-validation performed on the entire dataset. The final model was fitted for the sample of unrelated twins. We used the trained coefficients (i.e., regression weights) to identify the most predictive G and E factors. The joint effect of the polygenic scores and environmental variables was estimated by fitting all G and E predictors together in elastic net models for each developmental psychopathology phenotype and observing the additional variance explained.

#### Supplementary Note 5: Testing for gene environment correlation (rGE) using mediation models.

The mediation model estimates the indirect effect of the predictor (X) on the outcome (Y) via a mediator, i.e., an intervening variable (mediator; M) by regressing M on X and regressing Y on both X and M using two separate equations ^13,14^:

1) M_i_ = d_M_ + aX_i_+ e_M.i_

Where M_i_ is the mediator for individual i; d_M_ is the intercept for the mediator (M); _a_X_i_ is the slope of M regressed on the predictor (X) and e_M.i_ is the measurement error for individual i.

2) Y_i_ = d_Y_ + bM_i_ + cX_i_ + e_Y.i_
Where Y_i_ is the outcome for individual i; d_Y_ is the intercept for the outcome (Y); bM_i_ is the slope of the outcome (Y) regressed on the mediator (M) controlling for the predictor (X); cX_i_ is the slope of the outcome (Y) regressed on the predictor (X) controlling for the mediator (M) and e_Y.i_ is the measurement error for individual i.

The indirect effect of the predictor (X) on the outcome (Y) via the mediator (M) is calculated as:

$$\hat{a} \times\hat{b}$$

Where a and b are the estimated coefficients for the paths X 🡪 M and M 🡪 Y respectively.

In cases where:

$$\hat{c}= \hat{a} \times\hat{b}+ \hat{c}'$$

we can interpret the total effect of X on Yc’ as the sum of the indirect effect (a×b) and the direct effect (c’).

#### Supplementary Note 6: Distributions and data transformations.

Skewedness of environmental and developmental psychopathology measures was assessed based on histograms and the skew statistic. Variables were transformed based on the skew being lower than -1 or greater than 1. Supplementary Table 14 presents the transformation methods used and comparison of skews prior to and following the transformation. Supplementary Figures 8 and 9 show distributions of environmental and developmental psychopathology scales. Supplementary Figure 10 shows correlations between untransformed and transformed variables.

### Supplementary Tables

Supplementary Table 1. Results of the G models, predicting symptoms of internalising and externalising psychopathology in adolescence using polygenic scores.	7

Supplementary Table 2. Results of the E models, predicting symptoms of internalising and externalising psychopathology in adolescence using environmental measures.	8

Supplementary Table 3. Results of the G+E models, predicting symptoms of internalising and externalising psychopathology using polygenic scores and environmental measures.	9

Supplementary Table 4. Results of the G+E models, predicting symptoms of internalising and externalising psychopathology using polygenic scores and environmental measures in the total sample, including dizygotic co-twins.	10

Supplementary Table 5. Results of the cross-rater G+E models, predicting parent-rated symptoms of internalising and externalising psychopathology using polygenic scores and self-rated environmental measures and vice versa.	11

Supplementary Table 6. Results of the G+E models, predicting symptoms of internalising and externalising psychopathology using polygenic scores and environmental measures in male only sample.	12

Supplementary Table 7. Results of the G+E models, predicting symptoms of internalising and externalising psychopathology using polygenic scores and environmental measures in female only sample.	13

Supplementary Table 8. Results of the G+E+G×E models, predicting symptoms of internalising and externalising psychopathology using polygenic scores, environmental measures and their interaction.	14

Supplementary Table 9. Results of the mediation models testing for gene-environment correlation in symptoms of internalising and externalising psychopathology.	16

Supplementary Table 10. Representativeness of the selected sample.	17

Supplementary Table 11. List of the polygenic scores.	17

Supplementary Table 12. List of the environmental variables.	17

Supplementary Table 13. List of the environmental variables.	20

Supplementary Table 14. Transformation methods and skew statistics.	27

Supplementary Table 1. Results of the G models, predicting symptoms of internalising and externalising psychopathology in adolescence using polygenic scores.

| Psychopathology | R2 | RMSE | alpha | lambda | N |
| --- | --- | --- | --- | --- | --- |
| Parent-rated data | | | | | |
| SDQ total behaviour problems | 0.030 | 0.977 | 0.5 | 0.030 | 4010 |
| Conner's total ADHD | 0.031 | 0.988 | 0.5 | 0.020 | 4010 |
| ICUT callous-unemotional traits | 0.019 | 0.963 | 0.5 | 0.010 | 4000 |
| MFQ depression | 0.021 | 0.990 | 0.5 | 0.010 | 4013 |
| ARBQ anxiety | 0.019 | 0.994 | 0.5 | 0.020 | 4011 |
| Self-rated data | | | | | |
| SDQ total behaviour problems | 0.031 | 0.983 | 0.5 | 0.040 | 3997 |
| SWAN total ADHD | 0.043 | 0.977 | 0.5 | 0.010 | 902 |
| ICUT callous-unemotional traits | 0.037 | 0.977 | 0.5 | 0.040 | 899 |
| MFQ depression | 0.016 | 1.002 | 0.5 | 0.030 | 4002 |
| CASI anxiety | 0.020 | 0.989 | 0.5 | 0.020 | 4000 |

*Note*. G models= models using the polygenic scores to predict symptoms of symptoms of internalising and externalising psychopathology; R2= proportion of variance explained; RMSE= root mean square error; N= sample size.

Supplementary Table 2. Results of the E models, predicting symptoms of internalising and externalising psychopathology in adolescence using environmental measures.

| Psychopathology | R2 | RMSE | alpha | lambda | N |
| --- | --- | --- | --- | --- | --- |
| Age 9 | | | | | |
| Parent-rated data | | | | | |
| SDQ total behaviour problems | 0.144 | 0.890 | 0.5 | 0.051 | 1672 |
| Conner's total ADHD | 0.116 | 0.920 | 0.5 | 0.040 | 1670 |
| ICUT callous-unemotional traits | 0.104 | 0.906 | 0.5 | 0.061 | 1670 |
| MFQ depression | 0.056 | 0.959 | 0.5 | 0.030 | 1670 |
| ARBQ anxiety | 0.060 | 0.961 | 0.5 | 0.051 | 1672 |
| Self-rated data | | | | | |
| SDQ total behaviour problems | 0.071 | 0.946 | 0.5 | 0.061 | 1519 |
| SWAN total ADHD | 0.040 | 0.961 | 0.5 | 0.081 | 616 |
| ICUT callous-unemotional traits | 0.077 | 0.971 | 0.5 | 0.131 | 614 |
| MFQ depression | 0.033 | 0.998 | 0.5 | 0.081 | 1520 |
| CASI anxiety | 0.011 | 0.965 | 0.5 | 0.121 | 1520 |
| Age 12 | | | | | |
| Parent-rated data | | | | | |
| SDQ total behaviour problems | 0.184 | 0.887 | 0.5 | 0.030 | 3263 |
| Conner's total ADHD | 0.156 | 0.905 | 0.5 | 0.010 | 3264 |
| ICUT callous-unemotional traits | 0.128 | 0.904 | 0.5 | 0.010 | 3255 |
| MFQ depression | 0.057 | 0.966 | 0.5 | 0.030 | 3264 |
| ARBQ anxiety | 0.064 | 0.961 | 0.5 | 0.010 | 3266 |
| Self-rated data | | | | | |
| SDQ total behaviour problems | 0.078 | 0.946 | 0.5 | 0.051 | 3184 |
| SWAN total ADHD | 0.072 | 0.963 | 0.5 | 0.061 | 722 |
| ICUT callous-unemotional traits | 0.097 | 0.948 | 0.5 | 0.061 | 720 |
| MFQ depression | 0.041 | 0.979 | 0.5 | 0.030 | 3187 |
| CASI anxiety | 0.018 | 0.982 | 0.5 | 0.040 | 3189 |
| Age 16 | | | | | |
| Parent-rated data | | | | | |
| SDQ total behaviour problems | 0.049 | 0.924 | 0.5 | 0.030 | 1059 |
| Conner's total ADHD | 0.024 | 0.929 | 0.5 | 0.040 | 1059 |
| ICUT callous-unemotional traits | 0.034 | 0.921 | 0.5 | 0.071 | 1060 |
| MFQ depression | NA | 0.910 | 0.5 | 1.000 | 1060 |
| ARBQ anxiety | 0.020 | 0.959 | 0.5 | 0.000 | 1060 |
| Self-rated data | | | | | |
| SDQ total behaviour problems | 0.184 | 0.878 | 0.5 | 0.020 | 1526 |
| SWAN total ADHD | 0.106 | 0.895 | 0.5 | 0.071 | 618 |
| ICUT callous-unemotional traits | 0.175 | 0.881 | 0.5 | 0.061 | 615 |
| MFQ depression | 0.134 | 0.938 | 0.5 | 0.030 | 1531 |
| CASI anxiety | 0.073 | 0.927 | 0.5 | 0.020 | 1532 |

*Note*. E models= models using environmental data to predict symptoms of internalising and externalising psychopathology; R2= proportion of variance explained; RMSE= root mean square error; N= sample size.

Supplementary Table 3. Results of the G+E models, predicting symptoms of internalising and externalising psychopathology using polygenic scores and environmental measures.

| Psychopathology | R2 | RMSE | alpha | lambda | N |
| --- | --- | --- | --- | --- | --- |
| Age 9 | | | | | |
| Parent-rated data | | | | | |
| SDQ total behaviour problems | 0.164 | 0.879 | 0.5 | 0.030 | 1722 |
| Conner's total ADHD | 0.120 | 0.924 | 0.5 | 0.051 | 1720 |
| ICUT callous-unemotional traits | 0.122 | 0.900 | 0.5 | 0.040 | 1720 |
| MFQ depression | 0.074 | 0.951 | 0.5 | 0.030 | 1670 |
| ARBQ anxiety | 0.075 | 0.954 | 0.5 | 0.051 | 1672 |
| Self-rated data | | | | | |
| SDQ total behaviour problems | 0.097 | 0.928 | 0.5 | 0.020 | 1674 |
| SWAN total ADHD | 0.058 | 0.952 | 0.5 | 0.040 | 680 |
| ICUT callous-unemotional traits | 0.092 | 0.949 | 0.5 | 0.061 | 678 |
| MFQ depression | 0.039 | 0.996 | 0.5 | 0.061 | 1675 |
| CASI anxiety | 0.023 | 0.961 | 0.5 | 0.111 | 1520 |
| Age 12 | | | | | |
| Parent-rated data | | | | | |
| SDQ total behaviour problems | 0.180 | 0.888 | 0.5 | 0.030 | 3334 |
| Conner's total ADHD | 0.152 | 0.910 | 0.5 | 0.020 | 3335 |
| ICUT callous-unemotional traits | 0.130 | 0.903 | 0.5 | 0.000 | 3326 |
| MFQ depression | 0.066 | 0.964 | 0.5 | 0.020 | 3335 |
| ARBQ anxiety | 0.070 | 0.960 | 0.5 | 0.020 | 3337 |
| Self-rated data | | | | | |
| SDQ total behaviour problems | 0.097 | 0.942 | 0.5 | 0.030 | 3319 |
| SWAN total ADHD | 0.089 | 0.954 | 0.5 | 0.030 | 751 |
| ICUT callous-unemotional traits | 0.115 | 0.940 | 0.5 | 0.030 | 749 |
| MFQ depression | 0.053 | 0.976 | 0.5 | 0.020 | 3322 |
| CASI anxiety | 0.028 | 0.981 | 0.5 | 0.020 | 3324 |
| Age 16 | | | | | |
| Parent-rated data | | | | | |
| SDQ total behaviour problems | 0.075 | 0.912 | 0.5 | 0.040 | 1059 |
| Conner's total ADHD | 0.041 | 0.922 | 0.5 | 0.061 | 1059 |
| ICUT callous-unemotional traits | 0.051 | 0.913 | 0.5 | 0.051 | 1060 |
| MFQ depression | 0.042 | 0.892 | 0.5 | 0.030 | 1060 |
| ARBQ anxiety | 0.035 | 0.954 | 0.5 | 0.081 | 1060 |
| Self-rated data | | | | | |
| SDQ total behaviour problems | 0.169 | 0.883 | 0.5 | 0.030 | 1597 |
| SWAN total ADHD | 0.128 | 0.884 | 0.5 | 0.020 | 643 |
| ICUT callous-unemotional traits | 0.170 | 0.879 | 0.5 | 0.071 | 640 |
| MFQ depression | 0.119 | 0.950 | 0.5 | 0.051 | 1603 |
| CASI anxiety | 0.060 | 0.938 | 0.5 | 0.030 | 1604 |

*Note*. G+E models= models using both the polygenic scores and environmental data to predict symptoms of internalising and externalising psychopathology; R2= proportion of variance explained; RMSE= root mean square error; N= sample size.

Supplementary Table 4. Results of the G+E models, predicting symptoms of internalising and externalising psychopathology using polygenic scores and environmental measures in the total sample, including dizygotic co-twins.

| Psychopathology | R2 | RMSE | alpha | lambda | N |
| --- | --- | --- | --- | --- | --- |
| Age 9 | | | | | |
| Parent-rated data | | | | | |
| SDQ total behaviour problems | 0.162 | 0.900 | 0.5 | 0.030 | 2538 |
| Conner's total ADHD | 0.126 | 0.936 | 0.5 | 0.030 | 2533 |
| ICUT callous-unemotional traits | 0.126 | 0.911 | 0.5 | 0.020 | 2535 |
| MFQ depression | 0.060 | 0.954 | 0.5 | 0.020 | 2535 |
| ARBQ anxiety | 0.076 | 0.947 | 0.5 | 0.030 | 2536 |
| Self-rated data | | | | | |
| SDQ total behaviour problems | 0.085 | 0.941 | 0.5 | 0.040 | 2467 |
| SWAN total ADHD | 0.027 | 0.973 | 0.5 | 0.121 | 986 |
| ICUT callous-unemotional traits | 0.067 | 0.958 | 0.5 | 0.051 | 983 |
| MFQ depression | 0.042 | 0.995 | 0.5 | 0.040 | 2469 |
| CASI anxiety | 0.016 | 0.978 | 0.5 | 0.051 | 2469 |
| Age 12 | | | | | |
| Parent-rated data | | | | | |
| SDQ total behaviour problems | 0.175 | 0.896 | 0.5 | 0.020 | 5002 |
| Conner's total ADHD | 0.146 | 0.907 | 0.5 | 0.020 | 5001 |
| ICUT callous-unemotional traits | 0.124 | 0.912 | 0.5 | 0.030 | 4990 |
| MFQ depression | 0.064 | 0.963 | 0.5 | 0.010 | 5003 |
| ARBQ anxiety | 0.069 | 0.953 | 0.5 | 0.010 | 5004 |
| Self-rated data | | | | | |
| SDQ total behaviour problems | 0.093 | 0.942 | 0.5 | 0.020 | 4978 |
| SWAN total ADHD | 0.075 | 0.954 | 0.5 | 0.030 | 1088 |
| ICUT callous-unemotional traits | 0.081 | 0.940 | 0.5 | 0.051 | 1085 |
| MFQ depression | 0.051 | 0.979 | 0.5 | 0.020 | 4982 |
| CASI anxiety | 0.020 | 0.979 | 0.5 | 0.010 | 4983 |
| Age 16 | | | | | |
| Parent-rated data | | | | | |
| SDQ total behaviour problems | 0.072 | 0.931 | 0.5 | 0.030 | 1580 |
| Conner's total ADHD | 0.045 | 0.936 | 0.5 | 0.040 | 1580 |
| ICUT callous-unemotional traits | 0.051 | 0.920 | 0.5 | 0.020 | 1582 |
| MFQ depression | 0.029 | 0.904 | 0.5 | 0.040 | 1582 |
| ARBQ anxiety | 0.044 | 0.955 | 0.5 | 0.030 | 1582 |
| Self-rated data | | | | | |
| SDQ total behaviour problems | 0.171 | 0.882 | 0.5 | 0.020 | 2355 |
| SWAN total ADHD | 0.089 | 0.915 | 0.5 | 0.091 | 928 |
| ICUT callous-unemotional traits | 0.148 | 0.869 | 0.5 | 0.051 | 927 |
| MFQ depression | 0.105 | 0.957 | 0.5 | 0.030 | 2362 |
| CASI anxiety | 0.048 | 0.945 | 0.5 | 0.020 | 2363 |

*Note*. G+E models= models using both the polygenic scores and environmental data to predict symptoms of internalising and externalising psychopathology; R2= proportion of variance explained; RMSE= root mean square error; N= sample size.

Supplementary Table 5. Results of the cross-rater G+E models, predicting parent-rated symptoms of internalising and externalising psychopathology using polygenic scores and self-rated environmental measures and vice versa.

| Psychopathology | R2 | RMSE | alpha | lambda | N |
| --- | --- | --- | --- | --- | --- |
| Age 9 | | | | | |
| Parent-rated data | | | | | |
| SDQ total behaviour problems | 0.106 | 0.902 | 0.5 | 0.040 | 1692 |
| Conner's total ADHD | 0.072 | 0.938 | 0.5 | 0.030 | 1689 |
| ICUT callous-unemotional traits | 0.084 | 0.907 | 0.5 | 0.020 | 1689 |
| MFQ depression | 0.030 | 0.967 | 0.5 | 0.020 | 1689 |
| ARBQ anxiety | 0.032 | 0.965 | 0.5 | 0.030 | 1691 |
| Self-rated data | | | | | |
| SDQ total behaviour problems | 0.099 | 0.934 | 0.5 | 0.061 | 1703 |
| SWAN total ADHD | 0.092 | 0.944 | 0.5 | 0.071 | 694 |
| ICUT callous-unemotional traits | 0.068 | 0.967 | 0.5 | 0.111 | 692 |
| MFQ depression | 0.050 | 0.994 | 0.5 | 0.051 | 1704 |
| CASI anxiety | 0.023 | 0.969 | 0.5 | 0.091 | 1704 |
| Age 12 | | | | | |
| Parent-rated data | | | | | |
| SDQ total behaviour problems | 0.107 | 0.927 | 0.5 | 0.030 | 3331 |
| Conner's total ADHD | 0.085 | 0.945 | 0.5 | 0.020 | 3332 |
| ICUT callous-unemotional traits | 0.083 | 0.930 | 0.5 | 0.010 | 3323 |
| MFQ depression | 0.035 | 0.981 | 0.5 | 0.020 | 3332 |
| ARBQ anxiety | 0.035 | 0.976 | 0.5 | 0.010 | 3334 |
| Self-rated data | | | | | |
| SDQ total behaviour problems | 0.078 | 0.951 | 0.5 | 0.030 | 3323 |
| SWAN total ADHD | 0.081 | 0.961 | 0.5 | 0.020 | 753 |
| ICUT callous-unemotional traits | 0.084 | 0.956 | 0.5 | 0.051 | 751 |
| MFQ depression | 0.040 | 0.983 | 0.5 | 0.030 | 3326 |
| CASI anxiety | 0.024 | 0.984 | 0.5 | 0.010 | 3328 |
| Age 16 | | | | | |
| Parent-rated data | | | | | |
| SDQ total behaviour problems | 0.110 | 0.882 | 0.5 | 0.020 | 1598 |
| Conner's total ADHD | 0.085 | 0.916 | 0.5 | 0.040 | 1598 |
| ICUT callous-unemotional traits | 0.109 | 0.872 | 0.5 | 0.030 | 1596 |
| MFQ depression | 0.042 | 0.937 | 0.5 | 0.051 | 1597 |
| ARBQ anxiety | 0.049 | 0.950 | 0.5 | 0.020 | 1598 |
| Self-rated data | | | | | |
| SDQ total behaviour problems | 0.089 | 0.905 | 0.5 | 0.030 | 1054 |
| SWAN total ADHD | 0.064 | 0.934 | 0.5 | 0.081 | 442 |
| ICUT callous-unemotional traits | 0.041 | 0.953 | 0.5 | 0.111 | 440 |
| MFQ depression | 0.054 | 0.959 | 0.5 | 0.030 | 1060 |
| CASI anxiety | 0.033 | 0.964 | 0.5 | 0.071 | 1059 |

*Note*. G+E models= models using both the polygenic scores and environmental data to predict symptoms of internalising and externalising psychopathology; R2= proportion of variance explained; RMSE= root mean square error; N= sample size.

Supplementary Table 6. Results of the G+E models, predicting symptoms of internalising and externalising psychopathology using polygenic scores and environmental measures in male only sample.

| Psychopathology | R2 | RMSE | alpha | lambda | N |
| --- | --- | --- | --- | --- | --- |
| Age 9 | | | | | |
| Parent-rated data | | | | | |
| SDQ total behaviour problems | 0.203 | 0.835 | 0.5 | 0.071 | 735 |
| Conner's total ADHD | 0.162 | 0.938 | 0.5 | 0.081 | 736 |
| ICUT callous-unemotional traits | 0.170 | 0.911 | 0.5 | 0.040 | 736 |
| MFQ depression | 0.109 | 0.877 | 0.5 | 0.061 | 735 |
| ARBQ anxiety | 0.079 | 0.912 | 0.5 | 0.071 | 736 |
| Self-rated data | | | | | |
| SDQ total behaviour problems | 0.109 | 0.916 | 0.5 | 0.030 | 700 |
| SWAN total ADHD | 0.117 | 0.927 | 0.5 | 0.081 | 266 |
| ICUT callous-unemotional traits | 0.112 | 0.974 | 0.5 | 0.071 | 266 |
| MFQ depression | 0.053 | 0.913 | 0.5 | 0.071 | 700 |
| CASI anxiety | 0.033 | 0.955 | 0.5 | 0.061 | 700 |
| Age 12 | | | | | |
| Parent-rated data | | | | | |
| SDQ total behaviour problems | 0.181 | 0.873 | 0.5 | 0.040 | 1465 |
| Conner's total ADHD | 0.171 | 0.933 | 0.5 | 0.030 | 1468 |
| ICUT callous-unemotional traits | 0.126 | 0.941 | 0.5 | 0.051 | 1461 |
| MFQ depression | 0.093 | 0.892 | 0.5 | 0.020 | 1466 |
| ARBQ anxiety | 0.073 | 0.927 | 0.5 | 0.030 | 1467 |
| Self-rated data | | | | | |
| SDQ total behaviour problems | 0.095 | 0.912 | 0.5 | 0.030 | 1451 |
| SWAN total ADHD | 0.185 | 0.878 | 0.5 | 0.051 | 289 |
| ICUT callous-unemotional traits | 0.071 | 1.019 | 0.5 | 0.242 | 289 |
| MFQ depression | 0.052 | 0.898 | 0.5 | 0.040 | 1451 |
| CASI anxiety | 0.020 | 0.969 | 0.5 | 0.071 | 1453 |
| Age 16 | | | | | |
| Parent-rated data | | | | | |
| SDQ total behaviour problems | 0.063 | 0.924 | 0.5 | 0.121 | 420 |
| Conner's total ADHD | 0.029 | 0.957 | 0.5 | 0.182 | 420 |
| ICUT callous-unemotional traits | 0.037 | 0.965 | 0.5 | 0.152 | 420 |
| MFQ depression | 0.031 | 0.854 | 0.5 | 0.162 | 420 |
| ARBQ anxiety | 0.057 | 0.919 | 0.5 | 0.081 | 420 |
| Self-rated data | | | | | |
| SDQ total behaviour problems | 0.152 | 0.852 | 0.5 | 0.051 | 629 |
| SWAN total ADHD | 0.131 | 0.859 | 0.5 | 0.141 | 235 |
| ICUT callous-unemotional traits | 0.136 | 0.922 | 0.5 | 0.162 | 235 |
| MFQ depression | 0.107 | 0.877 | 0.5 | 0.061 | 632 |
| CASI anxiety | 0.051 | 0.925 | 0.5 | 0.061 | 633 |

*Note*. G+E models= models using both the polygenic scores and environmental data to predict symptoms of internalising and externalising psychopathology; R2= proportion of variance explained; RMSE= root mean square error; N= sample size.

Supplementary Table 7. Results of the G+E models, predicting symptoms of internalising and externalising psychopathology using polygenic scores and environmental measures in female only sample.

| Psychopathology | R2 | RMSE | alpha | lambda | N |
| --- | --- | --- | --- | --- | --- |
| Age 9 | | | | | |
| Parent-rated data | | | | | |
| SDQ total behaviour problems | 0.152 | 0.907 | 0.5 | 0.061 | 987 |
| Conner's total ADHD | 0.112 | 0.901 | 0.5 | 0.061 | 984 |
| ICUT callous-unemotional traits | 0.120 | 0.872 | 0.5 | 0.040 | 984 |
| MFQ depression | 0.055 | 1.006 | 0.5 | 0.061 | 985 |
| ARBQ anxiety | 0.092 | 0.973 | 0.5 | 0.071 | 986 |
| Self-rated data | | | | | |
| SDQ total behaviour problems | 0.104 | 0.929 | 0.5 | 0.030 | 974 |
| SWAN total ADHD | - | 0.979 | 0.5 | 1.000 | 414 |
| ICUT callous-unemotional traits | 0.118 | 0.916 | 0.5 | 0.081 | 412 |
| MFQ depression | 0.038 | 1.050 | 0.5 | 0.081 | 975 |
| CASI anxiety | 0.021 | 0.977 | 0.5 | 0.111 | 975 |
| Age 12 | | | | | |
| Parent-rated data | | | | | |
| SDQ total behaviour problems | 0.183 | 0.898 | 0.5 | 0.040 | 1869 |
| Conner's total ADHD | 0.141 | 0.888 | 0.5 | 0.020 | 1867 |
| ICUT callous-unemotional traits | 0.132 | 0.875 | 0.5 | 0.040 | 1865 |
| MFQ depression | 0.058 | 1.012 | 0.5 | 0.030 | 1869 |
| ARBQ anxiety | 0.076 | 0.980 | 0.5 | 0.030 | 1870 |
| Self-rated data | | | | | |
| SDQ total behaviour problems | 0.106 | 0.961 | 0.5 | 0.040 | 1868 |
| SWAN total ADHD | 0.077 | 0.980 | 0.5 | 0.071 | 462 |
| ICUT callous-unemotional traits | 0.123 | 0.908 | 0.5 | 0.081 | 460 |
| MFQ depression | 0.059 | 1.031 | 0.5 | 0.051 | 1871 |
| CASI anxiety | 0.036 | 0.992 | 0.5 | 0.051 | 1871 |
| Age 16 | | | | | |
| Parent-rated data | | | | | |
| SDQ total behaviour problems | 0.086 | 0.906 | 0.5 | 0.071 | 639 |
| Conner's total ADHD | 0.055 | 0.901 | 0.5 | 0.081 | 639 |
| ICUT callous-unemotional traits | 0.067 | 0.878 | 0.5 | 0.061 | 640 |
| MFQ depression | 0.047 | 0.919 | 0.5 | 0.040 | 640 |
| ARBQ anxiety | 0.057 | 0.957 | 0.5 | 0.081 | 640 |
| Self-rated data | | | | | |
| SDQ total behaviour problems | 0.190 | 0.897 | 0.5 | 0.030 | 968 |
| SWAN total ADHD | 0.127 | 0.908 | 0.5 | 0.101 | 408 |
| ICUT callous-unemotional traits | 0.208 | 0.847 | 0.5 | 0.061 | 405 |
| MFQ depression | 0.134 | 0.990 | 0.5 | 0.051 | 971 |
| CASI anxiety | 0.067 | 0.946 | 0.5 | 0.040 | 971 |

*Note*. G+E models= models using both the polygenic scores and environmental data to predict symptoms of internalising and externalising psychopathology; R2= proportion of variance explained; RMSE= root mean square error; N= sample size.

Supplementary Table 8. Results of the G+E+G×E models, predicting symptoms of internalising and externalising psychopathology using polygenic scores, environmental measures and their interaction.

| Psychopathology | R2 | RMSE | alpha | lambda | N |
| --- | --- | --- | --- | --- | --- |
| Age 9 | | | | | |
| Self-rated data | | | | | |
| MFQ depression | 0.044 | 0.993 | 0.5 | 0.50 | 1675 |
| CASI anxiety | 0.032 | 0.963 | 0.5 | 0.030 | 1675 |
| Age 12 | | | | | |
| Parent-rated data | | | | | |
| MFQ depression | 0.068 | 0.963 | 0.5 | 0.20 | 3335 |
| Self-rated data | | | | | |
| MFQ depression | 0.060 | 0.973 | 0.5 | 0.20 | 3322 |
| CASI anxiety | 0.035 | 0.978 | 0.5 | 0.020 | 3324 |
| Age 16 | | | | | |
| Self-rated data | | | | | |
| MFQ depression | 0.143 | 0.937 | 0.5 | 0.20 | 1603 |
| CASI anxiety | 0.074 | 0.931 | 0.5 | 0.20 | 1604 |

*Note*. G+E+G×E models= models using the polygenic scores, environmental data and their interaction to predict symptoms of internalising and externalising psychopathology; R2= proportion of variance explained; RMSE= root mean square error; N= sample size.

Supplementary Table 9. Results of the mediation models testing for gene-environment correlation in symptoms of internalising and externalising psychopathology.

| Predictor | Outcome | Mediator | Indirect effect | SE | Z-value | P-value | Total | %total mediated |
| --- | --- | --- | --- | --- | --- | --- | --- | --- |
| Age 9 environments | | | | | | | | |
| Environmentally mediated effects | | | | | | | | |
| G-predicted values | Self-reported MFQ depression | E-predicted values | 0.179 | 0.047 | 3.819 | <0.001 | 1.224 | 14.6 |
| G-predicted values | Self-reported CASI anxiety | E-predicted values | 0.065 | 0.027 | 2.392 | 0.017 | 1.176 | 5.5 |
| Genetically confounded effects | | | | | | | | |
| E-predicted values | Self-reported MFQ depression | G-predicted values | 0.060 | 0.021 | 2.890 | 0.004 | 1.00 | 6.0 |
| E-predicted values | Self-reported CASI anxiety | G-predicted values | 0.094 | 0.035 | 2.660 | 0.008 | 1.00 | 9.4 |
| Age 12 environments | | | | | | | | |
| Environmentally mediated effects | | | | | | | | |
| G-predicted values | Self-reported MFQ depression | E-predicted values | 0.141 | 0.028 | 4.980 | <0.001 | 1.178 | 12.0 |
| G-predicted values | Self-reported CASI anxiety | E-predicted values | 0.061 | 0.020 | 3.019 | 0.003 | 1.157 | 5.3 |
| G-predicted values | Parent-reported MFQ depression | E-predicted values | 0.187 | 0.035 | 5.361 | <0.001 | 1.258 | 14.9 |
| Genetically confounded effects | | | | | | | | |
| E-predicted values | Self-reported MFQ depression | G-predicted values | 0.068 | 0.015 | 4.528 | <0.001 | 1.036 | 6.6 |
| E-predicted values | Self-reported CASI anxiety | G-predicted values | 0.072 | 0.024 | 3.067 | 0.002 | 1.197 | 6.0 |
| E-predicted values | Parent-reported MFQ depression | G-predicted values | 0.060 | 0.013 | 4.616 | <0.001 | 1.063 | 5.6 |
| Age 16 environments | | | | | | | | |
| Environmentally mediated effects | | | | | | | | |
| G-predicted values | Self-reported MFQ depression | E-predicted values | 0.330 | 0.069 | 4.795 | <0.001 | 1.199 | 27.5 |
| G-predicted values | Self-reported CASI anxiety | E-predicted values | 0.165 | 0.051 | 3.223 | 0.001 | 1.137 | 14.5 |
| Genetically confounded effects | | | | | | | | |
| E-predicted values | Self-reported MFQ depression | G-predicted values | 0.041 | 0.012 | 3.364 | 0.001 | 1.00 | 4.1 |
| E-predicted values | Self-reported CASI anxiety | G-predicted values | 0.042 | 0.016 | 2.708 | 0.007 | 1.00 | 4.2 |

*Note*. SE= standard error.

Supplementary Table 10. Representativeness of the selected sample.

| Ethnicity and SES | Selected sample | 1st Contact sample | National equivalents^a,b^ |
| --- | --- | --- | --- |
| % white | 99.9% | 91.7% | 93% |
| % mother A-levels or higher | 39.0% | 35.5% | 35% |
| % father A-levels or higher | 42.3% | 44.8% | 47% |
| % mother employed | 45.3% | 43.1% | 50% |
| % father employed | 85.8% | 91.6% | 91% |
| *Note*. ^a^ including cohort of parents with children born in late 1990s and early 2000s;  ^b^ derived from ^15.^ | | | |

Supplementary Table 11. List of the polygenic scores.

| Genome-wide association study (GWAS) | N (cases/controls) |
| --- | --- |
| ADHD^16^ | 8,691/38,691 |
| Alcohol dependence ^17^ | 10,206/28,480 |
| Anorexia nervosa ^18^ | 16,992/55,525 |
| Anxiety disorders ^19^ | 26,104/58,113 |
| ASD ^20^ | 18,382/27,969 |
| Bipolar disorder ^21^ | 41,917/ 371,549 |
| Major depressive disorder ^22^ | 170,756/329,443 |
| Externalising behaviour ^23^ | 1,492,085 |
| Neuroticism ^24^ | 390,821 |
| Obsessive-compulsive disorder ^25^ | 2,688/7,037 |
| Post-traumatic stress disorder ^26^ | 9,831/19,225 |
| Schizophrenia ^27^ | 39,910/60,558 |
| Tourette’s syndrome ^28^ | 4,819/9,488 |

*Note*. We used polygenic scores derived from GWAS of those disorders included in ^29^, unless newer GWAS have become available at the time of conducting analyses.

Supplementary Table 12. List of the environmental variables.

| Age of collection | Rater | Items & scales |
| --- | --- | --- |
| Age 9 | Parent | Parent Feelings scale ^30^   - Being a Parent: wish child would leave alone - Being a Parent: does not feel amused by child - Being a Parent: child makes me angry - Being a Parent: does not feel close to child - Being a Parent: feel frustrated by child - Being a Parent: not happy about relationship with child - Being a Parent: feel impatient with child   Parent Chaos scale ^31^   - Chaos: no regular bedtime routine - Chaos: cannot hear yourself think - Chaos: a real zoo - Chaos: we do not stay on top of things - Chaos: usually a TV on - Chaos: no calm atmosphere   Parent Discipline scale ^32^   - Discipline: does not explain or reason - Discipline: be firm or calm - Discipline: shout or tell off - Discipline: smack   Life events   - Life Event: Birth of Younger Sibling - Life Event: Divorce/Separation of Parents - Life Event: Death of Grandparent - Life Event: Death of Other Relative/Friend - Life Event: Financial Difficulties - Life Event: Hospitalisation of Elder Twin - Life Event: Hospitalisation of Parent - Life Event: Hospitalisation of Sibling - Life Event: Hospitalisation of Younger Twin - Life Event: Illness/Injury of Relative/Friend - Life Event: Moved House - Life Event: New Child - Life Event: New Parent Figure - Life Event: Other - Life Event: Prolonged Separation from Parent - Count of reported life events |
| Age 9 | Self | Self-reported Feelings scale ^30^   - Being a Parent: parent wishes I would leave alone - Being a Parent: parent does not find me funny - Being a Parent: I make parent angry - Being a Parent: I do not feel close to parent - Being a Parent: I make parent frustrated - Being a Parent: not happy about relationship with parent - Being a Parent: parent gets impatient   Self-reported Chaos scale ^31^   - Chaos: no regular bedtime routine - Chaos: cannot hear yourself think - Chaos: a real zoo - Chaos: we do not stay on top of things - Chaos: usually a TV on - Chaos: no calm atmosphere   Self-reported Discipline scale ^32^   - Discipline: parent rarely explains - Discipline: parents are not firm - Discipline: told off or shouted at - Discipline: smacked |
| Age 12 | Parent | Parent Feelings scale ^30^   - Parental Feelings: impatient - Parental Feelings: unhappy - Parental Feelings: not amused - Parental Feelings: leave me alone - Parental Feelings: angry - Parental Feelings: not close - Parental Feelings: frustrated   Parent Chaos scale ^31^   - Chaos: no regular bedtime routine - Chaos: cannot hear yourself think - Chaos: we do not stay on top of things - Chaos: usually a TV on - Chaos: no calm atmosphere   Parent Discipline scale ^32^   - Discipline: smack - Discipline: shout - Discipline: rarely explain - Discipline: not firm or calm |
| Age 12 | Self | Self-reported Feelings scale ^30^   - Being a Parent: parent gets impatient - Being a Parent: not happy about relationship with parent - Being a Parent: parent does not find me funny - Being a Parent: parent wishes I would leave alone - Being a Parent: I make parent angry - Being a Parent: I do not feel close to parent - Being a Parent: I make parent frustrated,   Self-reported Chaos scale ^31^   - Chaos: no regular bedtime routine - Chaos: cannot hear yourself think - Chaos: a real zoo - Chaos: we do not stay on top of things - Chaos: usually a TV on - Chaos: no calm atmosphere   Self-reported Discipline scale ^32^   - Discipline: smacked - Discipline: told off or shouted at - Discipline: parent rarely explains - Discipline: parents are not firm |
| Age 16 | Parent | - Father highest qualification level - Father SOC level - Mother highest qualification level - Mother SOC level - Household income level |
| Age 16 | Self | Self-reported Chaos scale ^31^   - Chaos: no regular routine - Chaos: cannot hear yourself think - Chaos: a real zoo - Chaos: we do not stay on top of things - Chaos: usually a TV on - Chaos: no calm atmosphere   Parental Control ^33^   - Parental Control: how late to stay up - Parental Control: which friends - Parental Control: which activities - Parental Control: meet friends - Parental Control: how you dress - Parental Control: what you do with your money - Parental Control: watch on TV - Parental Control: religious training   Parental Monitoring ^33^   - Parental Monitoring: who spend time with - Parental Monitoring: how spend free time - Parental Monitoring: how spend money - Parental Monitoring: where after school - Parental Monitoring: where on weekend - Parental Monitoring: problems at school |

Supplementary Table 13. List of the environmental variables.

| Age of collection | Rater | Items & scales |
| --- | --- | --- |
| Age 16 | Parent | Strengths and Difficulties Questionnaire (SDQ) ^1^   - My child: is restless, overactive, and cannot stay still for long - My child: has a hot temper - My child: generally follows instructions, and usually does what adults request - My child: is constantly fidgeting - My child: often fights with his/her friends and/or bullies them - My child: is easily distracted, concentration wanders - My child: thinks things out before acting - My child: steals from home, school, work or elsewhere - My child: sees tasks through to the end, has good attention span - My child: is considerate of other people’s feelings - My child: shares readily with others - My child: helpful if someone is hurt, upset, or feeling ill - My child: has at least one good friend - My child: is generally liked by others - My child: is kind to people younger than them - My child: often volunteers to help others (parents, teachers, peers, colleagues)   Conners Rating Scale ^2^   - My child: is always “on the go” or acts as if driven by a motor - My child: avoids, expresses reluctance about, or has difficulties engaging in tasks that require sustained mental effort - My child: has difficulties sustaining attention in tasks or activities - My child: does not seem to listen to what is being said to them - My child: runs about excessively in situations where it is inappropriate to do so - My child: does not follow through on instructions and fails to finish work, schoolwork, or chores - My child: has difficulty organizing tasks and activities - My child: talks excessively - My child: fails to give close attention to details or makes careless mistakes in work, schoolwork, or other activities - My child: has difficulty queuing or awaiting their turn in games or group situations - My child: interrupts conversations - My child: is forgetful in daily activities - My child: fidgets with hands or feet, squirms in their seat - My child: has difficulty playing or engaging in leisure activities quietly - My child: often loses things necessary for tasks or activities (e.g., work or schoolwork) - My child: regularly leaves their seat in the classroom or in other situations in which remaining seated is expected - My child: is easily distracted by things going on around them - My child: blurts out answers to questions before the questions have been completed - My child: changes mood quickly and drastically - My child: cries often and easily - My child: has temper outbursts – their behaviour can be explosive or unpredictable   Inventory of Callous-Unemotional Traits (ICUT) ^3^   - My child: expresses their feelings openly - My child: does not seem to know right from wrong - My child: cares about their work or schoolwork - My child: does not care who they hurt to get what they want - My child: feels bad or guilty when they have done something wrong - My child: does not show emotions - My child: does not care about being on time - My child: is concerned about the feelings of others - My child: cares if they are in trouble - My child: does not let feelings control them - My child: does not care about doing things well - My child: seems very cold and uncaring - My child: easily admits to being wrong - My child: makes it easy to tell how they are feeling - My child: always tries their best - My child: apologizes to people they have hurt - My child: tries not to hurt others' feelings - My child: shows no remorse when they have done something wrong - My child: is very expressive and emotional - My child: does not like to put time into doing things well - My child: finds the feelings of others unimportant - My child: hides their feelings from others - My child: works hard on everything - My child: does things to make others feel good   Anxiety-Related Behaviours Questionnaire (ARBQ) ^4^   - My child: is afraid of small, enclosed spaces, heights, water, or the dark - My child: takes a long time to warm to strangers - My child: is afraid in social situations - My child: tends to check things are done exactly right - My child: asks for reassurance that he/she is OK - My child: insists on doing something over and over, to the extent that it interferes with day-to-day life - My child: tends to be shy and timid - My child: is afraid of medical procedures such as going to see the doctor/dentist - My child: has twitches, mannerisms, or tics of the face and body - My child: doesn't tend to enjoy him/herself - My child: often makes comments critical of him/herself - My child: complains or whines a lot - My child: has low self-confidence - My child: is fussy or over-particular - My child: tends to blame him/herself - My child: is often extremely upset or distressed when a parent leaves, wound up or stressed - My child: often seems worked up, on edge, or tense - My child: is afraid of animals or insects (like dogs, spiders, or snakes) - My child: is anxious that bad things will happen   Mood and Feelings Questionnaire (MFQ) ^5^   - My child: didn’t enjoy anything at all - My child: felt unreasonably tired, so that he/she just sat around and did nothing - My child: felt he/she was no good anymore - My child: cried a lot - My child: found it hard to think properly or concentrate - My child: hated him/herself - My child: felt he/she was a bad person - My child: felt lonely - My child: thought nobody really loved him/her - My child: thought he/she could never be as good as other kids - My child: felt he/she did everything wrong |
| Age 16 | Self | Strengths and Difficulties Questionnaire (SDQ) ^1^   - I try to be nice to other people and care about their feelings - I am restless and cannot stay still for long - I get a lot of headaches, stomach-aches, or sickness - I usually share with others, such as food, games, or pens - I get very angry and often lose my temper - I am usually on my own and generally play alone or keep to myself - I usually do as I am told - I worry a lot - I am helpful if someone is hurt, upset, or feeling ill - I am constantly fidgeting or squirming - I have one good friend or more - I fight a lot and can make other people do what I want - Other people my age generally like me - I am easily distracted and find it difficult to concentrate - I am nervous in new situations and easily lose confidence - I am kind to younger children - I am often accused of lying or cheating - Other children or young people pick on me or bully me - I often volunteer to help others, including parents, teachers, and children - I think before I do things - I take things that are not mine from home, school, or elsewhere - I get on better with adults than with people my own age - I have many fears and am easily scared - I finish the work I’m doing, and my attention is good   Strengths and Weaknesses of Attention-Deficit/Hyperactivity Symptoms and Normal Behaviors (SWAN) ^6^   - I pay close attention to detail and avoid careless mistakes - I sustain attention on tasks or leisure activities - I listen when spoken to directly - I follow through on instructions and finish schoolwork or chores. I am organized in my tasks and activities - I engage in tasks that require sustained mental effort - I keep track of things necessary for activities - I ignore distractions that go on around me - I remember to do daily activities - I sit still and control the movement of my hands and feet - I stay seated when required to - I stop myself from moving about when it is inappropriate to do so - When engaging in leisure activities, I keep noise levels reasonable - I can settle down and rest, controlling constant activity - I am able to control how much I talk - I reflect on questions and control blurting out answers - I await my turn rather than queue jumping - I enter into conversations without interrupting   Inventory of Callous-Unemotional Traits (ICUT) ^3^   - I express my feelings openly - What I think is ‘right’ and ‘wrong’ is different from what other people think - I care about how well I do at school or work - I do not care who I hurt to get what I want - I feel bad or guilty when I do something wrong - I do not show my emotions to others - I do not care about being on time - I am concerned about the feelings of others - I do not care if I get into trouble - I do not let my feelings control me - I do not care about doing things well - I seem very cold and uncaring to others - I easily admit to being wrong - It is easy for others to tell how I am feeling - I always try my best - I apologize to someone if I hurt them - I try not to hurt others’ feelings - I do not feel remorseful when I do something wrong - I am very expressive and emotional - I do not like to put the time into doing things well - The feelings of others are unimportant to me - I hide my feelings from others - I work hard on everything I do - I do things to make others feel good   Childhood Anxiety Sensitivity Index (CASI) ^7^   - I don't want other people to know when I feel afraid - I worry that I might be going crazy when I cannot keep my mind on my schoolwork - It scares me when I feel "shaky" - It scares me when I feel like I am going to faint - It is important for me to stay in control of my feelings - It scares me when my heart beats fast - I feel embarrassed when my stomach rumbles or makes noise - It scares me when I feel like I am going to throw up - When I notice that my heart is beating fast, I worry that there might be something wrong with me - It scares me when I have trouble getting my breath - When my stomach hurts, I worry that I might be really ill - It scares me when I cannot concentrate on my schoolwork - Others my age can tell when I feel shaky - Unusual feelings in my body scare me - When I am afraid, I worry that I might be crazy - I get scared when I feel nervous - I don't like to let my feelings show - Funny feelings in my body scare me   Mood and Feelings Questionnaire (MFQ) ^5^   - I felt miserable or unhappy - I didn’t enjoy anything at all - I felt so tired I just sat around and did nothing - I was very restless - I felt I was no good anymore - I cried a lot - I found it hard to think properly or concentrate - I hated myself - I felt I was a bad person - I felt lonely - I thought that nobody really loved me - I thought I could never be as good as others - I did everything wrong |

Supplementary Table 14. Transformation methods and skew statistics.

| Variable | Rater | Transformation method | Skew before transformation | Skew after transformation |
| --- | --- | --- | --- | --- |
| Environmental data | | | | |
| Life events | Parent | Square root | 1.31 | 0.16 |
| Developmental psychopathology | | | | |
| SDQ total behaviour problems | Parent | Square root | 1.34 | -0.16 |
| Conners total ADHD | Parent | Square root | 2.04 | 0.34 |
| ARBQ anxiety | Parent | Square root | 2.10 | 0.36 |
| CASI anxiety | Self | Square root | 1.17 | -0.02 |
| MFQ depression | Parent | Inverse transformation | 4.00 | -1.02 |
| MFQ depression | Self | Square root | 1.90 | 0.41 |

### Supplementary Figures

Supplementary Figure 1. Elastic net coefficients of the association between parent-rated environments measures at ages 9, 12 and 16 and developmental psychopathology measures. Note. For clarity of visualisation, only those environments with coefficients lower than -0.01 or greater than 0.01 are included in the figure.	28

Supplementary Figure 2. Elastic net coefficients of the association between self-rated environments measures at ages 9, 12 and 16 and developmental psychopathology measures. Note. For clarity of visualisation, only those environments with coefficients lower than -0.01 or greater than 0.01 are included in the figure.	29

Supplementary Figure 3. Comparison of the proportion of variance explained by sensitivity G+E models.	30

Supplementary Figure 4. G×E that emerged as significant in the continuous analysis plotted for twins selected in the +/- 1 standard deviation quadrants of the polygenic scores and environments measured at age 9.	31

Supplementary Figure 5. G×E that emerged as significant in the continuous analysis plotted for twins selected in the +/- 1 standard deviation quadrants of the polygenic scores and environments measured at age 12.	32

Supplementary Figure 6. G×E that emerged as significant in the continuous analysis plotted for twins selected in the +/- 1 standard deviation quadrants of the polygenic scores and environments measured at age 16.	33

Supplementary Figure 7. Correlations between predicted values from G and E models for parent- and self-reported symptoms of psychopathology	34

Supplementary Figure 8. Distributions of environmental data.	35

Supplementary Figure 9. Distributions of psychopathology measures.	36

Supplementary Figure 10. Correlations between untransformed and transformed variables.	37

Supplementary Figure 11. Mediation and gene environment correlation (rGE) models. Panel A presents the mediation model of X on Y, mediated by M. Panel B presents the rGE model of G on behaviour problems, mediated by E. Panel C presents the rGE model of genetic confounding. Circles indicate residuals. Parameters a, b and c represent regression weights. Parameters x, y and z represent variance parameters. Note. Model illustrated in panel C is abstract due to the fact that G cannot by causally influenced by E.	38

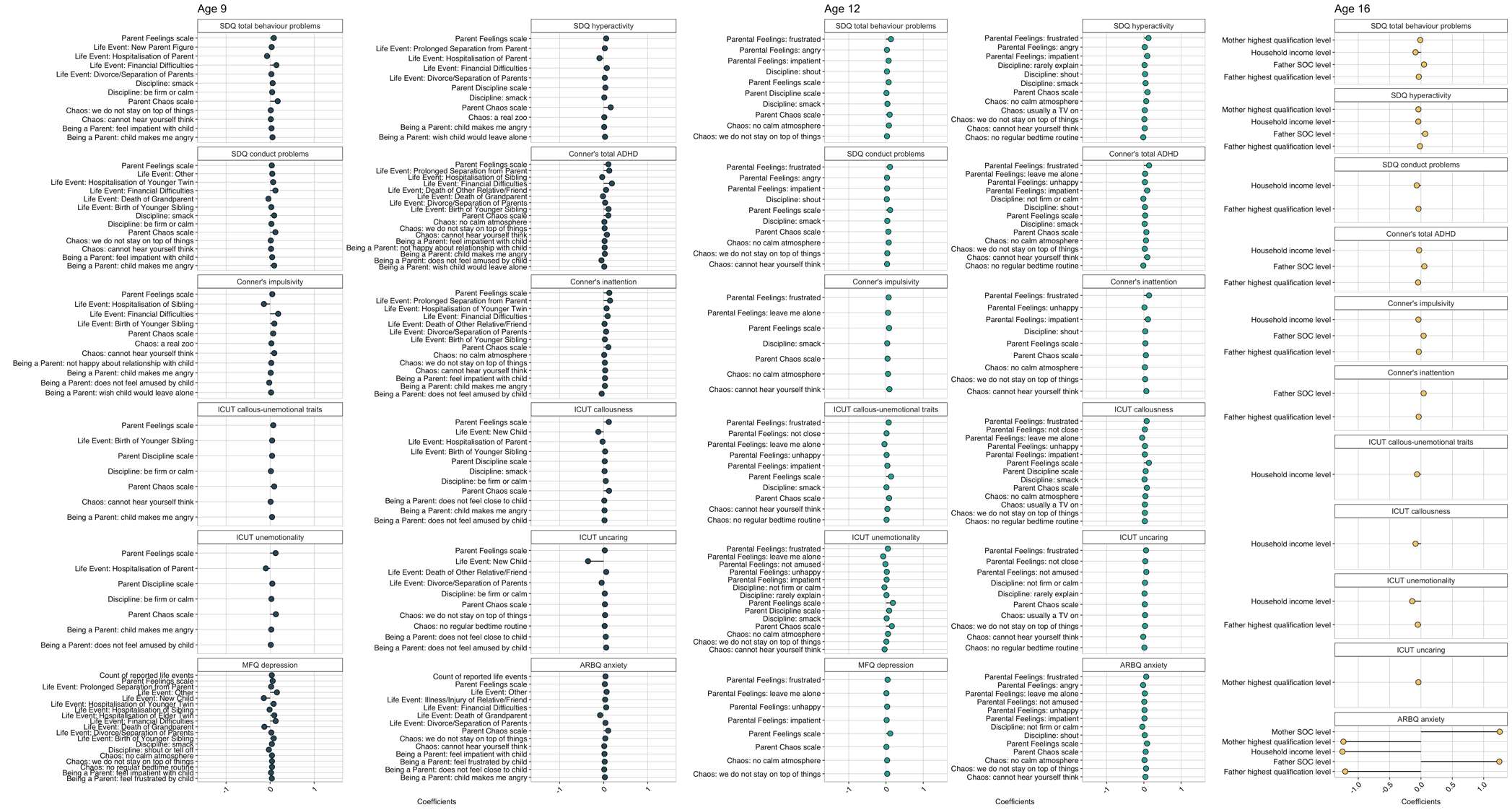

Supplementary Figure 1. Elastic net coefficients of the association between parent-rated environments measures at ages 9, 12 and 16 and developmental psychopathology measures. Note. For clarity of visualisation, only those environments with coefficients lower than -0.01 or greater than 0.01 are included in the figure.

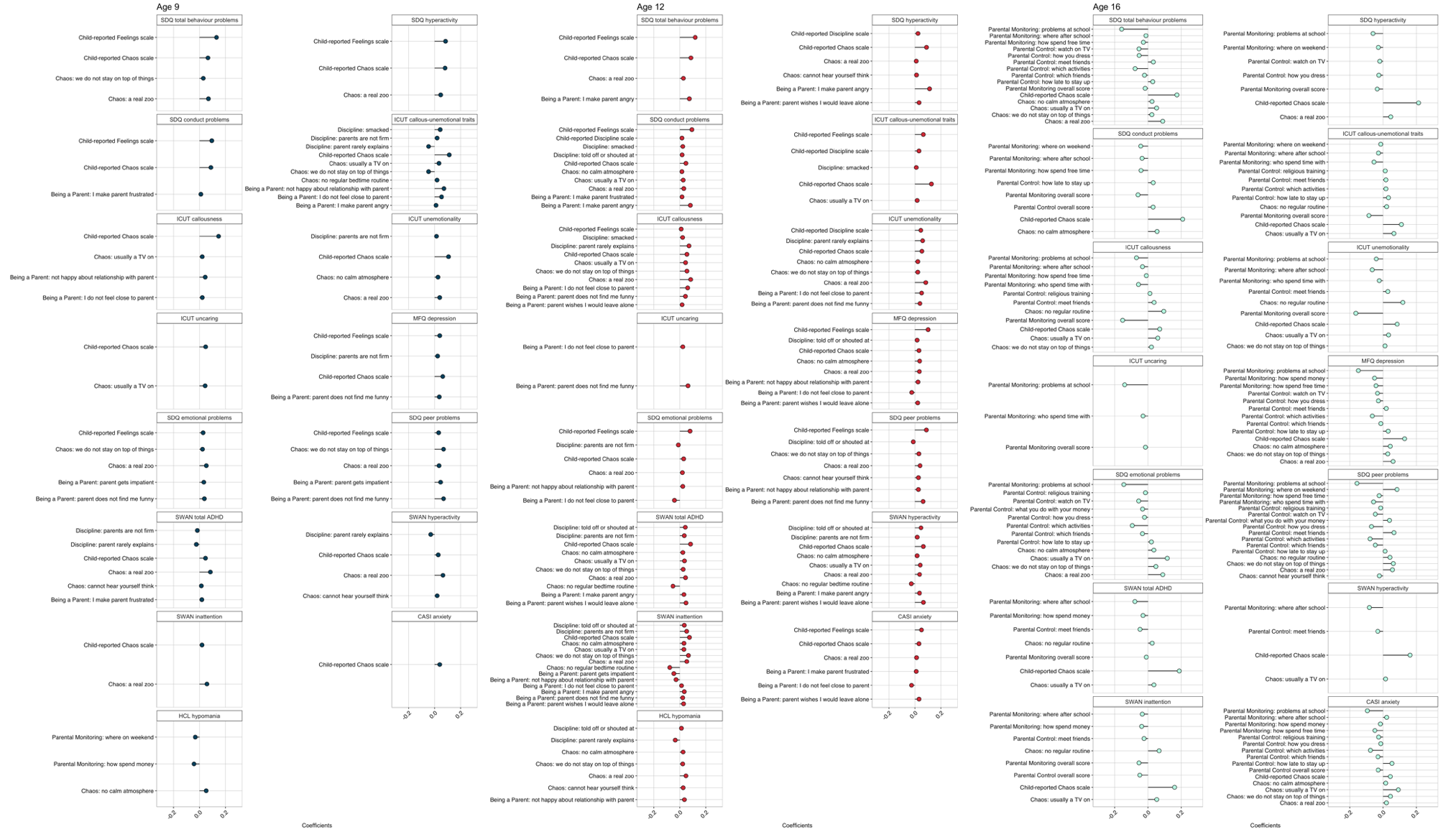

Supplementary Figure 2. Elastic net coefficients of the association between self-rated environments measures at ages 9, 12 and 16 and developmental psychopathology measures. Note. For clarity of visualisation, only those environments with coefficients lower than -0.01 or greater than 0.01 are included in the figure.

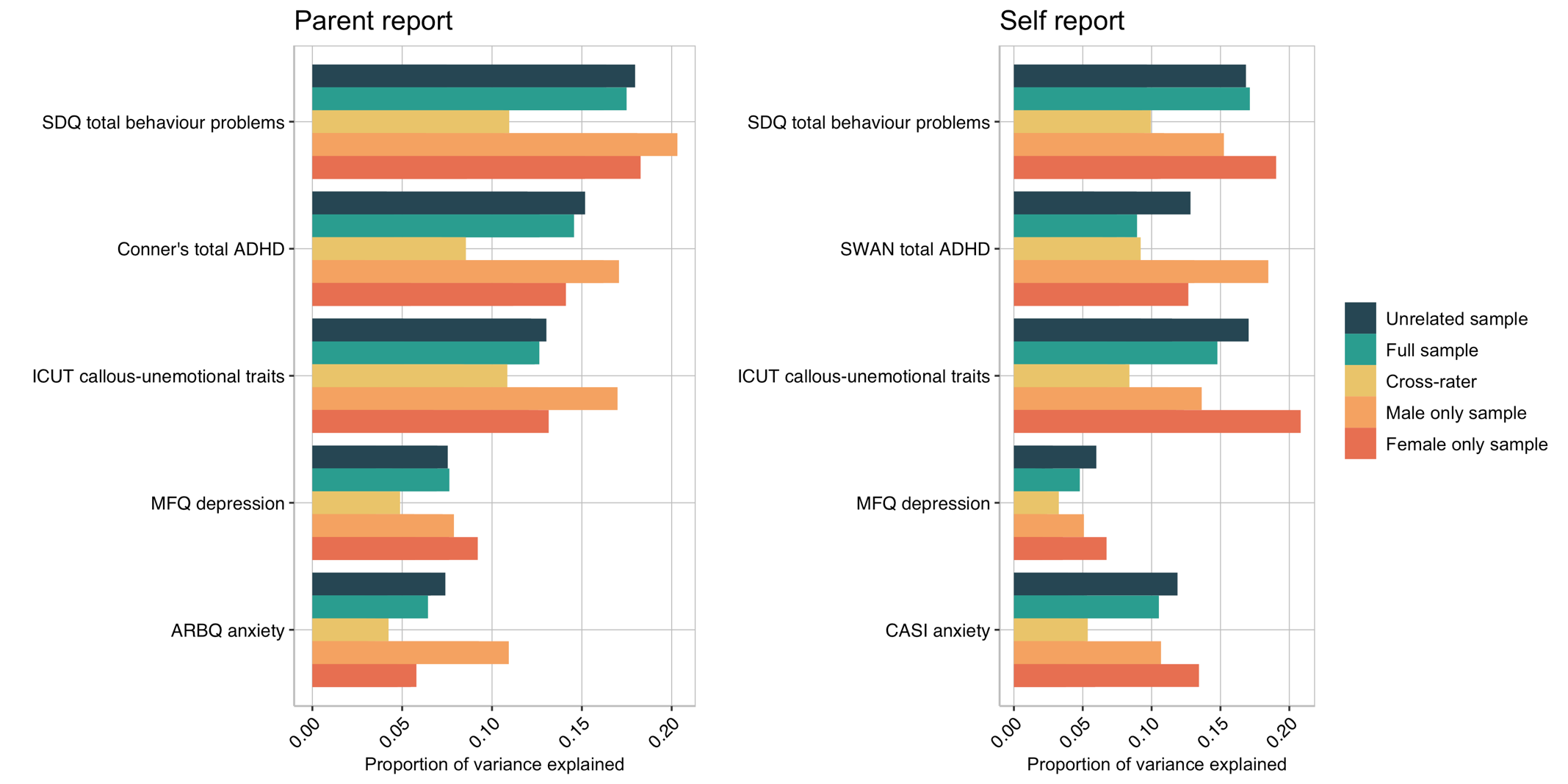

Supplementary Figure 3. Comparison of the proportion of variance explained by sensitivity G+E models.

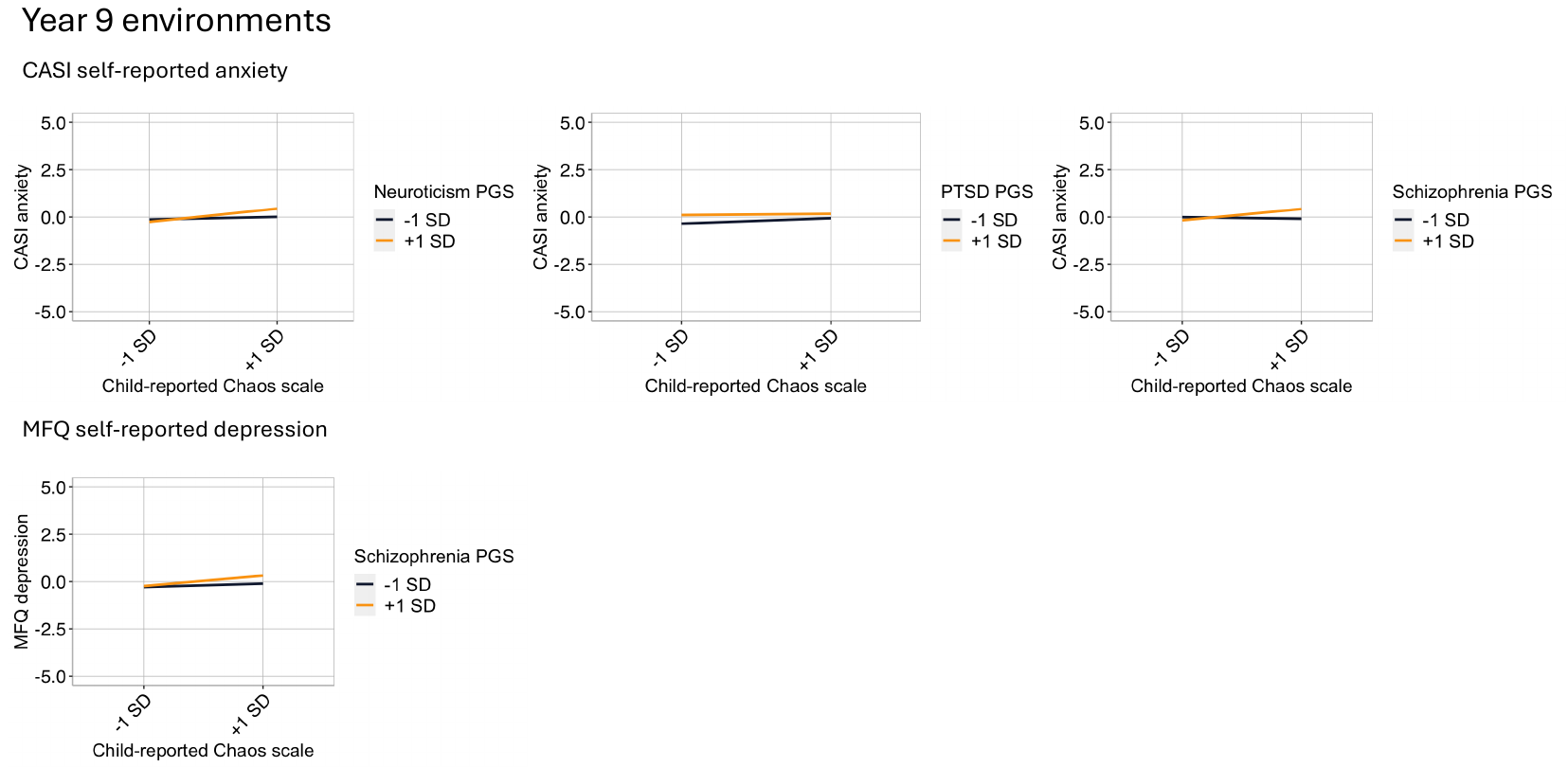

Supplementary Figure 4. G×E that emerged as significant in the continuous analysis plotted for twins selected in the +/- 1 standard deviation quadrants of the polygenic scores and environments measured at age 9.

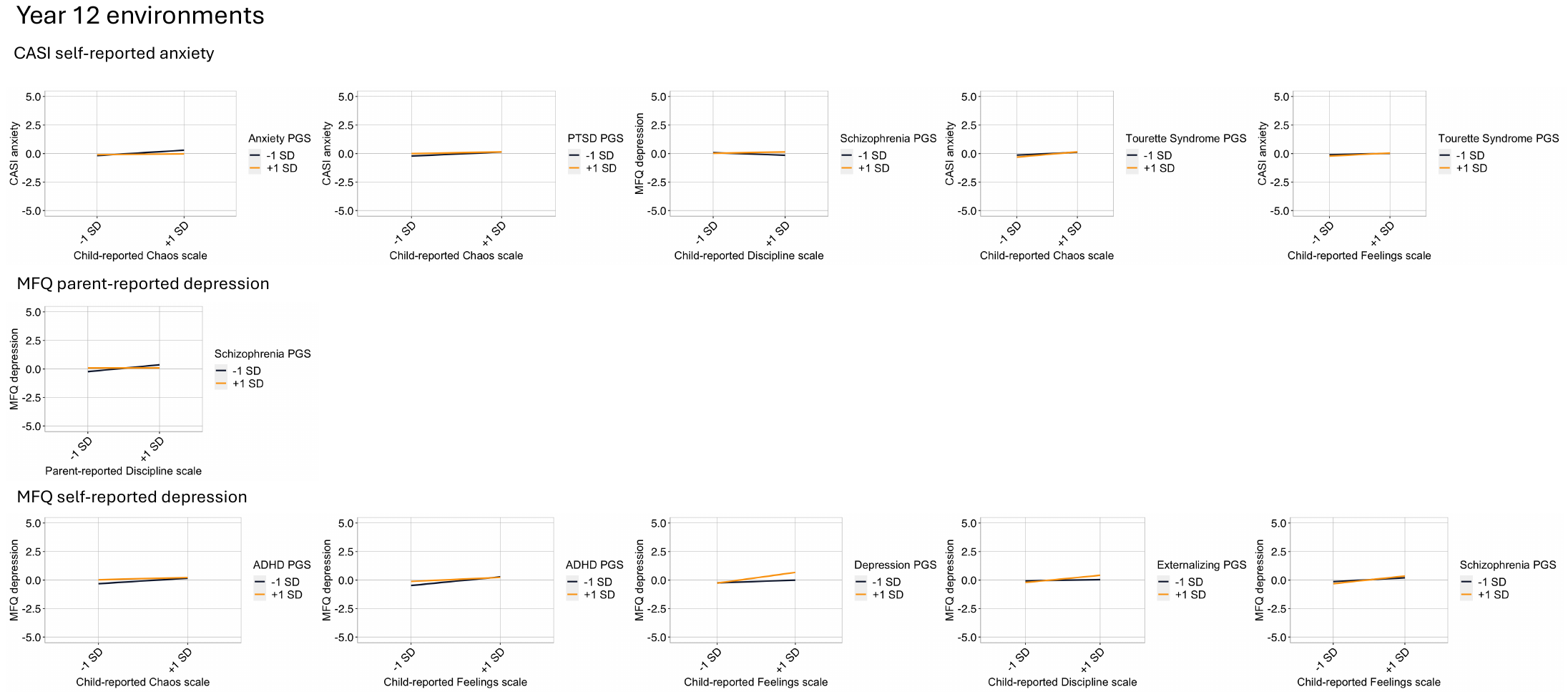

Supplementary Figure 5. G×E that emerged as significant in the continuous analysis plotted for twins selected in the +/- 1 standard deviation quadrants of the polygenic scores and environments measured at age 12.

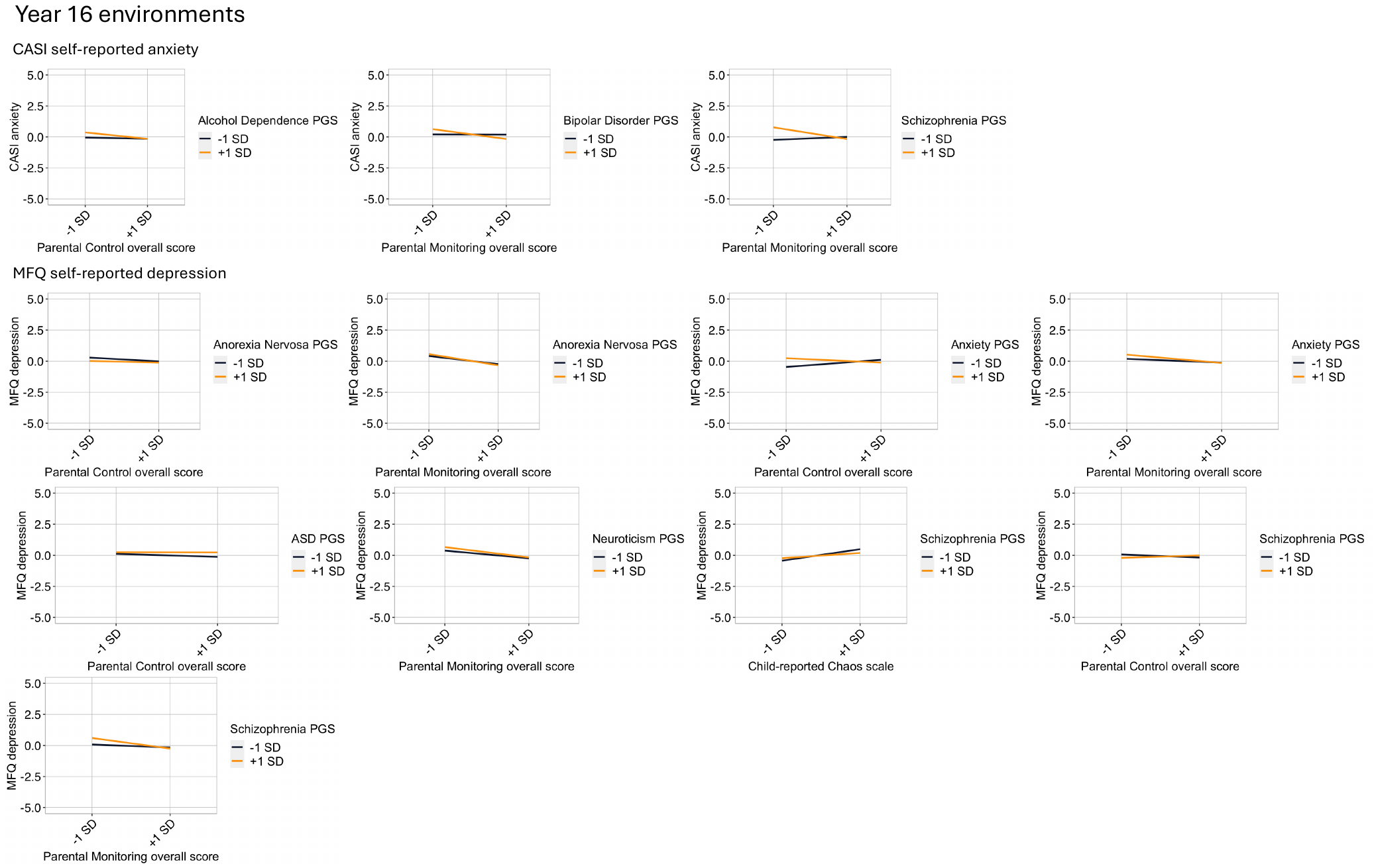

Supplementary Figure 6. G×E that emerged as significant in the continuous analysis plotted for twins selected in the +/- 1 standard deviation quadrants of the polygenic scores and environments measured at age 16.

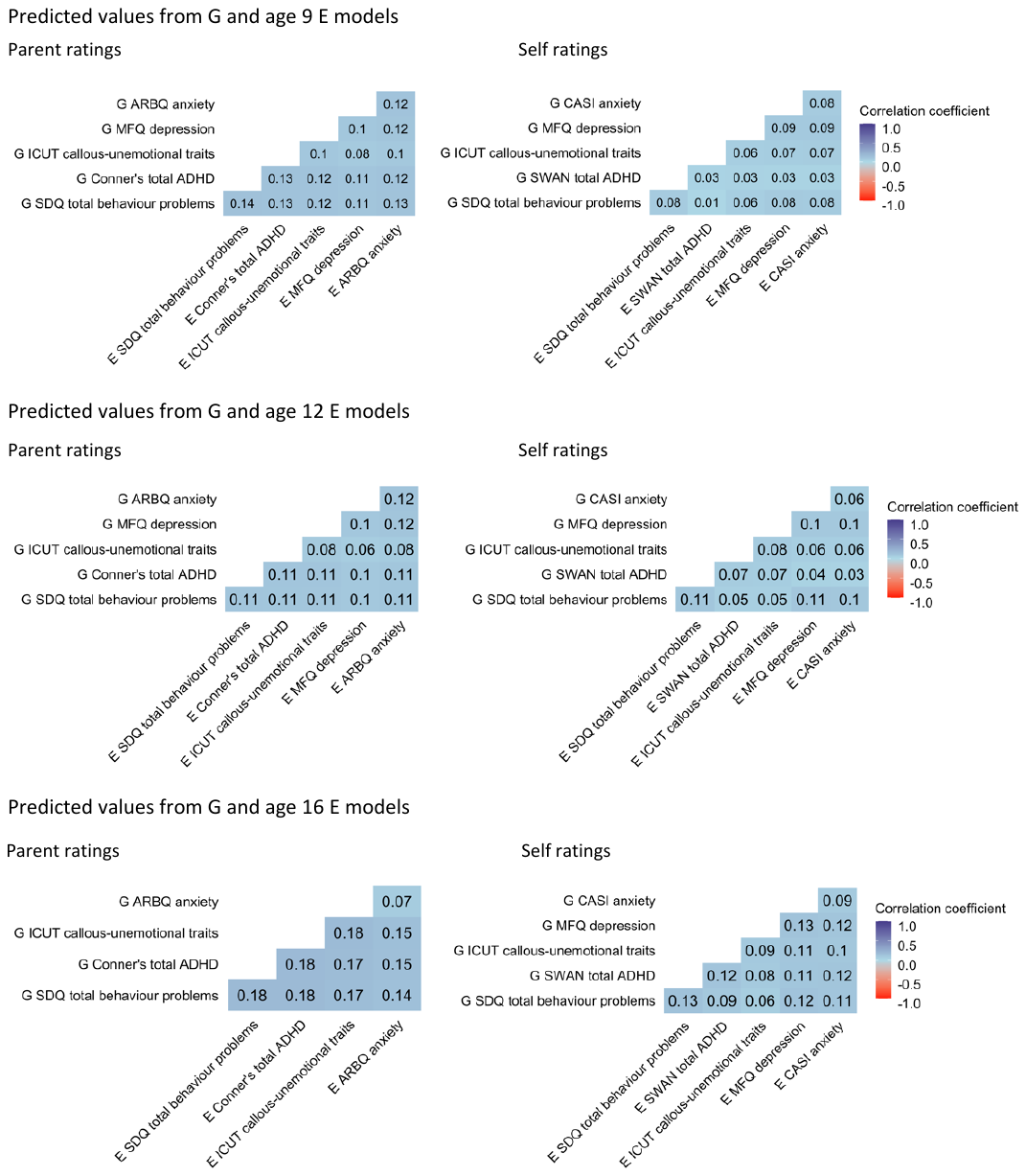

Supplementary Figure 7. Correlations between predicted values from G and E models for parent- and self-reported symptoms of psychopathology

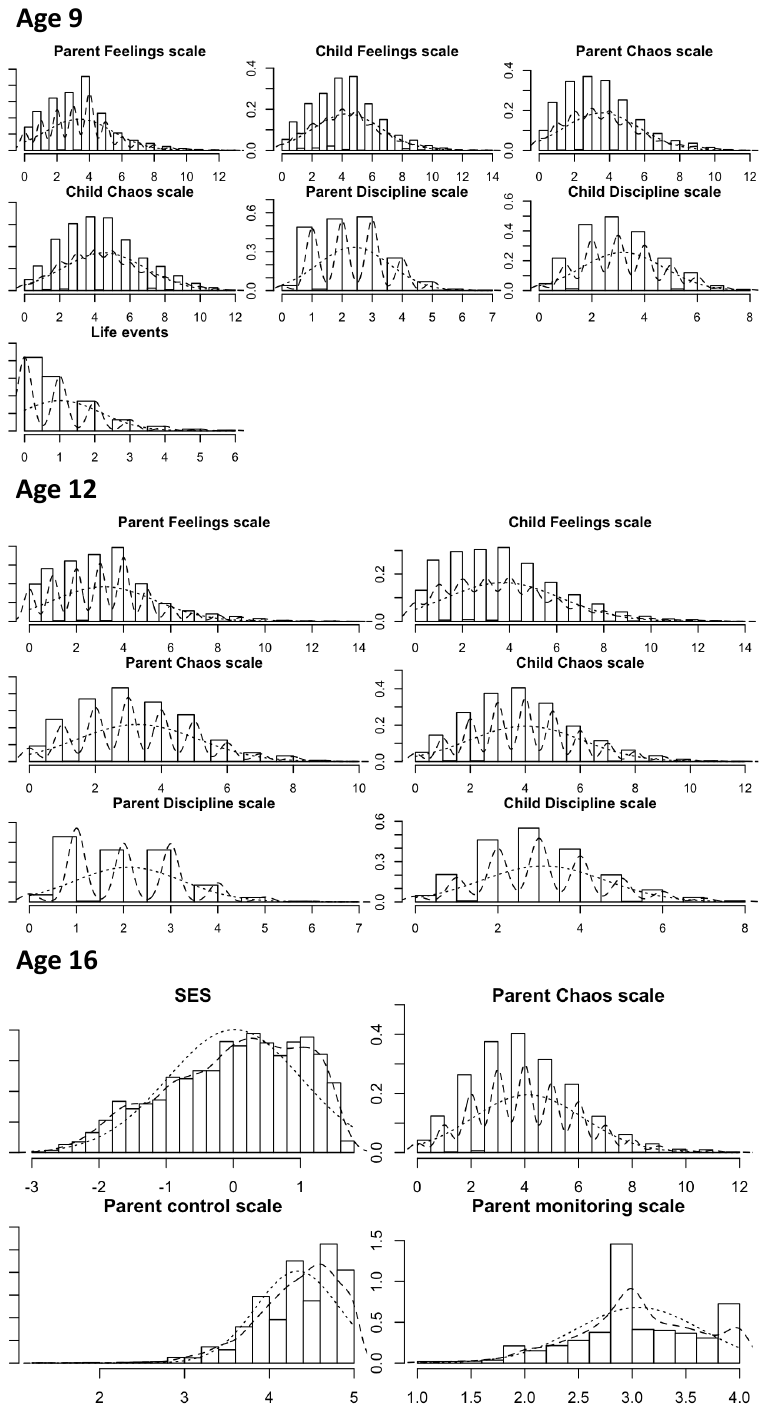

Supplementary Figure 8. Distributions of environmental data.

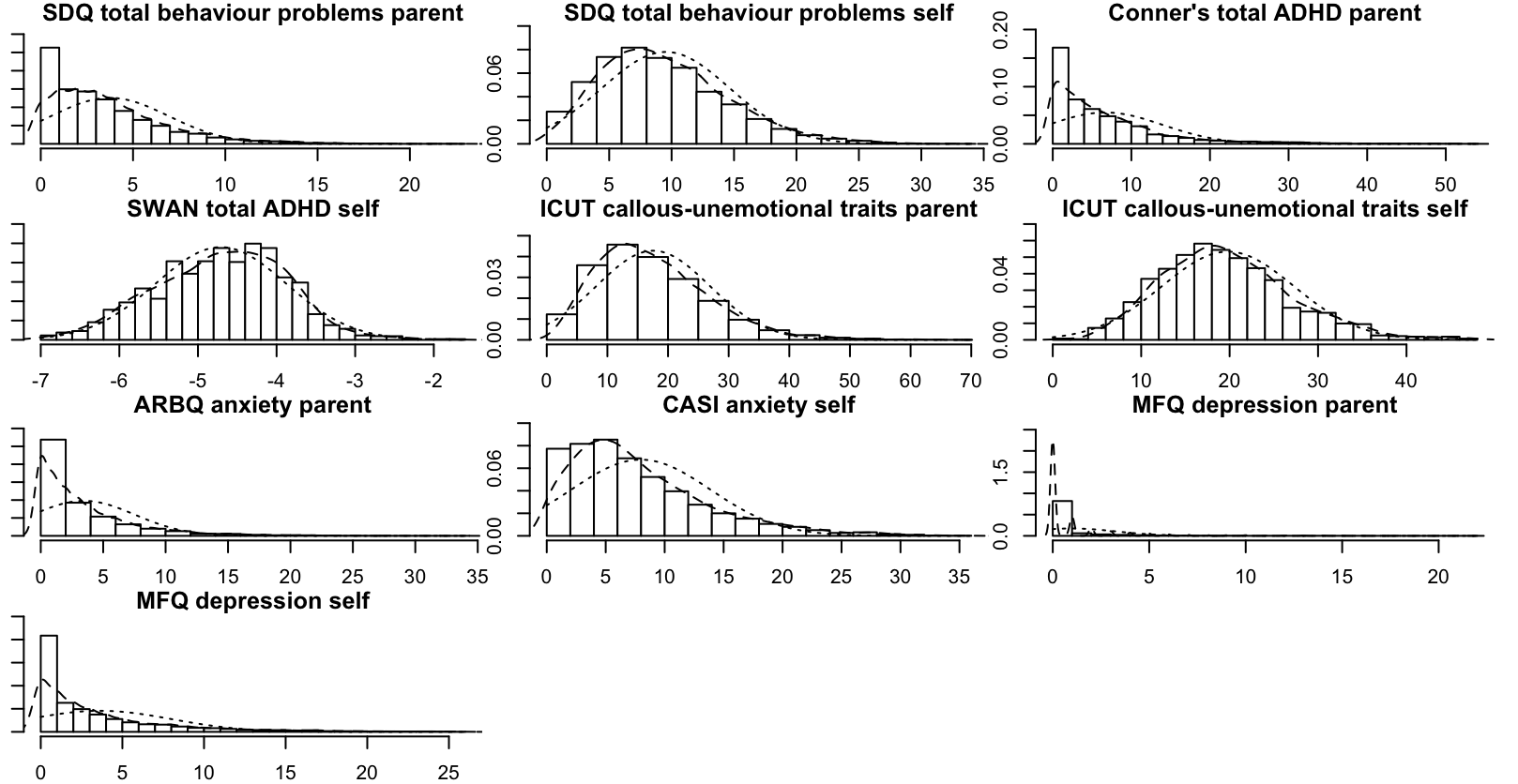

Supplementary Figure 9. Distributions of psychopathology measures.

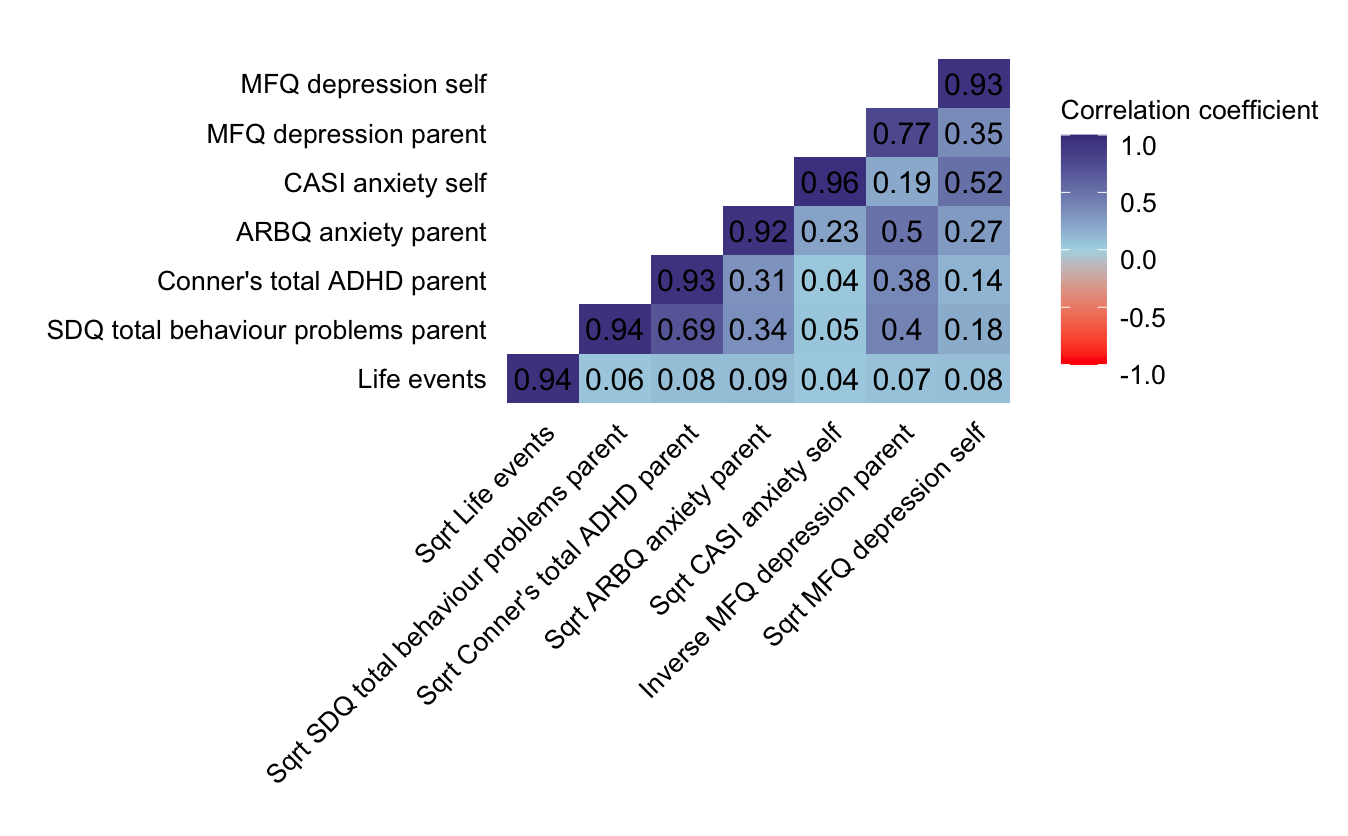

Supplementary Figure 10. Correlations between untransformed and transformed variables.

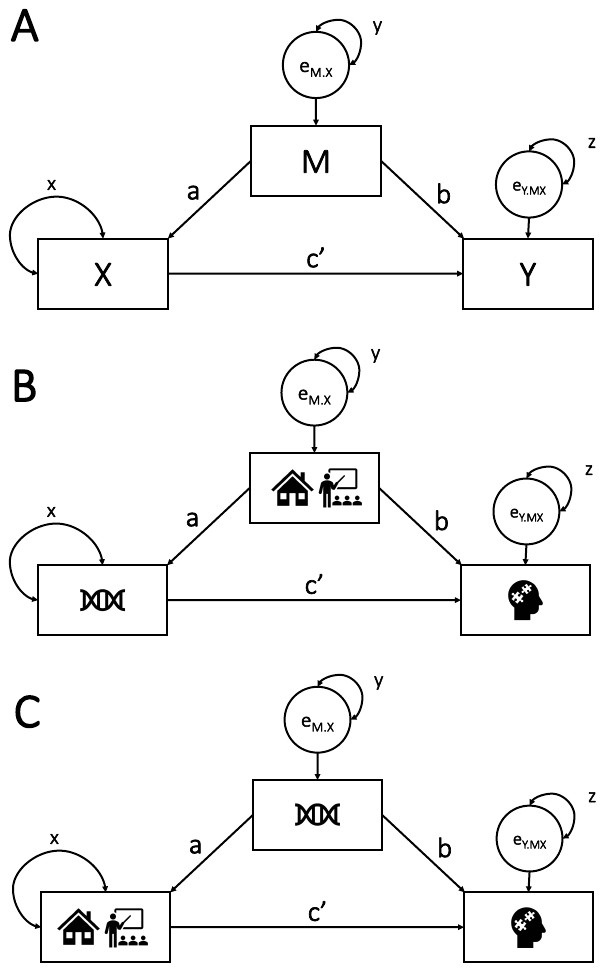

Supplementary Figure 11. Mediation and gene environment correlation (rGE) models. Panel A presents the mediation model of X on Y, mediated by M. Panel B presents the rGE model of G on behaviour problems, mediated by E. Panel C presents the rGE model of genetic confounding. Circles indicate residuals. Parameters a, b and c represent regression weights. Parameters x, y and z represent variance parameters. Note. Model illustrated in panel C is abstract due to the fact that G cannot by causally influenced by E.
